# Retrospective, observational analysis of the first one hundred consecutive cases of personalized bacteriophage therapy of difficult-to-treat infections facilitated by a Belgian consortium

**DOI:** 10.1101/2023.08.28.23294728

**Authors:** Jean-Paul Pirnay, Sarah Djebara, Griet Steurs, Johann Griselain, Christel Cochez, Steven De Soir, Tea Glonti, An Spiessens, Emily Vanden Berghe, Sabrina Green, Jeroen Wagemans, Cédric Lood, Eddie Schrevens, Nina Chanishvili, Mzia Kutateladze, Mathieu de Jode, Pieter-Jan Ceyssens, Jean-Pierre Draye, Gilbert Verbeken, Daniel De Vos, Thomas Rose, Jolien Onsea, Brieuc Van Nieuwenhuyse, Bacteriophage Therapy Providers, Bacteriophage Donors, Patrick Soentjens, Rob Lavigne, Maya Merabishvili

## Abstract

In contrast to the many reports of successful cases of personalized bacteriophage therapy, randomized controlled trials of non-personalized bacteriophage products did not bring the expected results. Here, we present the outcomes of a retrospective, observational analysis of the first 100 consecutive cases of personalized bacteriophage therapy of difficult-to-treat infections facilitated by a Belgian consortium. The most common indications were lower respiratory tract, skin & soft tissue, and bone infections, and involved combinations of 26 bacteriophages, individually selected and sometimes pre-adapted to target the causative bacterial pathogens. Clinical improvement and eradication of the targeted bacteria were reported for 77.2% and 61.3% of infections, respectively. Eradication was 70% less probable when no concomitant antibiotics were used (odds-ratio = 0.3; 95% confidence interval = 0.127–0.749). In vivo selection of bacteriophage resistance and in vitro bacteriophage-antibiotic synergy were documented in 43.8% (7/16 patients) and 90% (9/10) of evaluated patients, respectively. Bacteriophage immune neutralization was observed in 38.5% (5/13) of screened patients. (BT100 study, ClinicalTrials.gov registration: NCT05498363.)

---

Antimicrobial resistance (AMR) is a prominent global health threat with an estimated 1.27 million attributable deaths in 2019^1^, and only a few small companies attempting to develop and market new classes of antibiotics in an environment lacking commercial sustainability. Even though the recent launch of an AMR Action to support research and development is expected to bring up to four new antibiotics to the market by 2030^2^, there is an urgent need to seek alternative antimicrobial strategies. Bacteriophage therapy (BT), the use of bacteriophages – the viruses of bacteria – to treat bacterial infections, was first applied by Félix d’Hérelle in 1919^3^. This antibacterial therapy immediately experienced a heyday in the West, which lasted until the large-scale production and supply of antibiotics during World War II. Thereafter, BT quickly fell into decline, only to be further developed and applied in the former Soviet Union and its spheres of influence, which were initially shielded from antibiotic development. Some twenty years ago, this traditional therapy was rediscovered by Western scientists and entrepreneurs, and hailed as a possible savior to the accelerating AMR crisis^4^.

A recent systematic review confirmed that BT can generally be considered as safe, with a low incidence of adverse events, and could be a promising strategy in the fight against AMR^5^. The authors, however, acknowledged that high-quality trials are urgently required to build scientific knowledge and make useful predictions on the outcome of bacteriophage treatments. A number of companies are currently attempting to develop and market defined broad-spectrum BT products in compliance with contemporary requirements, which involves good manufacturing practices (GMP) certification, preclinical research (toxicity and pharmacology), and conducting randomized controlled trials (RCTs). However, the handful of bacteriophage RCTs that have been performed up to date, have not brought the expected results in terms of effectiveness^6^. A commonly reported reason for these disappointing results is the use of invariable one-size-fits-all bacteriophage products^6^.

In contrast, an increasing number of successful BT cases are reported in the scientific literature^5^. Irrespective of an obvious positive result publication bias, it is striking that in most of these successful cases tailored bacteriophage products were used. In addition, these personalized bacteriophage preparations, which were shown to target the infecting bacteria *in vitro* prior to their clinical application, were often used in combination with antibiotics. When appropriate, bacteriophage preparations were even adapted to counter bacterial resistance that had emerged against the applied bacteriophages during BT^7^, or bacteriophages were pre-adapted (“trained”)^8^, or even engineered^9^ to be more effective. While general purpose and tailored BT products are complementary and should both prevail, the long and expensive conventional drug development and marketing pathways form insurmountable obstacles for personalized BT concepts^10^. Meanwhile, there are no BT products on the market in the Western world, and ad hoc, emergency solutions are being set up at national levels to enable the use of personalized BT in difficult-to-treat infections^11^.

Even though small series of similar BT cases are increasingly being reported^12–18^, the accumulation of singular BT case reports has resulted in a considerable heterogeneity in terms of bacteriophage product design, quality and titer, and BT protocols (e.g. route and frequency of administration and treatment duration). Since 2008, BT is used in Belgium – under the umbrella of Article 37 (Unproven Interventions in Clinical Practice) of the Declaration of Helsinki – as an additional tool in the fight against antimicrobial resistance. In 2018, Belgium implemented a BT framework, which centers on the magistral preparation (compounding pharmacies in the US) of tailor-made bacteriophage medicines^19^, and is increasingly finding appeal in other European countries^11^. Within this framework, prospective clinical studies can be run using standardized protocols^20^, while also allowing for deviating (from the study protocols) and urgent cases to be treated.

A Belgian BT consortium, consisting of the Queen Astrid military hospital (QAMH), KU Leuven, and Sciensano (formerly known as the Scientific Institute of Public Health) facilitated BT in about 140 difficult-to-treat infections in patients in Belgium and abroad (as of July 2023), not taking into account the patients treated in the context of prospective clinical trials. The selection of patients was largely based on clinical need, regulatory approval, and the availability of well-characterized bacteriophages targeting the infecting bacteria (Extended Data Fig. 1). Of note, the vast majority of selected cases concerned personalized BT as salvage therapy after standard antibiotic treatments had failed. Personalized bacteriophage preparations were produced at the QAMH, in accordance with the rules in force in the territory at the time of their use in clinical practice. Quality and safety of the bacteriophage preparations were verified by Sciensano. The BT protocols that were suggested to the treating physicians were based on the experiences of the George Eliava Institute of Bacteriophages, Microbiology and Virology (Eliava Institute) in Tbilisi, Republic of Georgia (personal communications), and on the application instructions of the Ministries of Health and of Medical and Microbiology Industry of the former USSR^21–23^.

Three supporting assays were offered to the treating physicians, without obligation, to allow for improved BT management: (i) Monitoring of the *in vivo* emergence of bacteriophage resistance using sequential bacterial samples isolated during BT; (ii) Analysis of the in vitro bacteriophage-antibiotic interactions prior to the start of BT; (iii) Evaluation of bacteriophage immune neutralization, or the ability of the patient’s serum to neutralize therapeutic bacteriophages. Demographic and clinical data were collected through a medical form. Clinical improvement (or not), eradication of the targeted bacterium (or not), and the advent, severity and duration of adverse events or reactions were assessed by the treating physicians.

In this article, we report the retrospective, observational analysis of the first 100 consecutive BT cases of difficult-to-treat infections, enabled by this Belgian consortium. Because all cases were included in this study, and not only successful, interesting or challenging cases, we were able to evaluate how often personalized BT, as offered by our consortium, produced a positive clinical outcome (general efficacy) and to identify functional relationships that are general in all cases. The knowledge gained from these cases will help physicians to select effective treatment protocols and to design new clinical trials.

## Results

### Patients and infections

Personalized BT of 100 consecutive patients targeted 114 difficult-to-treat infections (as diagnosed by the treating physicians), including 14 co-infections. Supplementary Table 1 provides an overview of these BT cases, which were performed by in total 63 Bacteriophage Therapy Providers* in 35 hospitals, 29 cities, and 12 countries (Fig. 1a). Twenty-seven of the 100 BT cases/patients were previously reported^8,16,24–36^. Since 2008, the number of BT cases performed under the umbrella of different regulatory frameworks, and facilitated by the Belgian consortium, has increased steadily (Fig. 1b). The prevalence of the main infection types is shown in Fig. 1c. The most common indications for BT include lower respiratory tract infections (LRTI, 25.4%, (29/114 infections)), skin & soft tissue infections (SSTI, 22.8% (26/114)), bone infections (BoneI, 14.0% (16/114)), and upper respiratory tract infections (URTI, 11.4% (13/114)). Fourteen patients presented with a co-infection, more specifically a blood stream infection (BSI, *n* = 10), a urinary tract infection (UTI, *n* = 2), an SSTI (*n* = 1), or an URTI (*n* = 1). Age and gender distribution are shown in Fig. 1d. The median age of the patients was 53 years (1–91 years), and 56.7% of the atients were male. Of note, five patients were one year or younger. Fourteen bacterial species were targeted (Fig. 1e), with the highest prevalence for *Pseudomonas aeruginosa* (44/100 patients) and *Staphylococcus aureus* (35/100 patients). Bacteriophages were administered intravenously to 20 patients (Supplementary Table 1); in ten of them as stand-alone BT, in ten concomitantly with intralesional (*n* = 4), nebulized (*n* = 3), topical (*n* = 2), or generalized (multiple application routes; *n* = 1) bacteriophage application. In ten patients, intravenous bacteriophages were used to treat or prevent blood stream infections. In 69.3% (79/114) of targeted infections, bacteriophages were administered in combination with standard-of-care antibiotics.

**Fig. 1.**
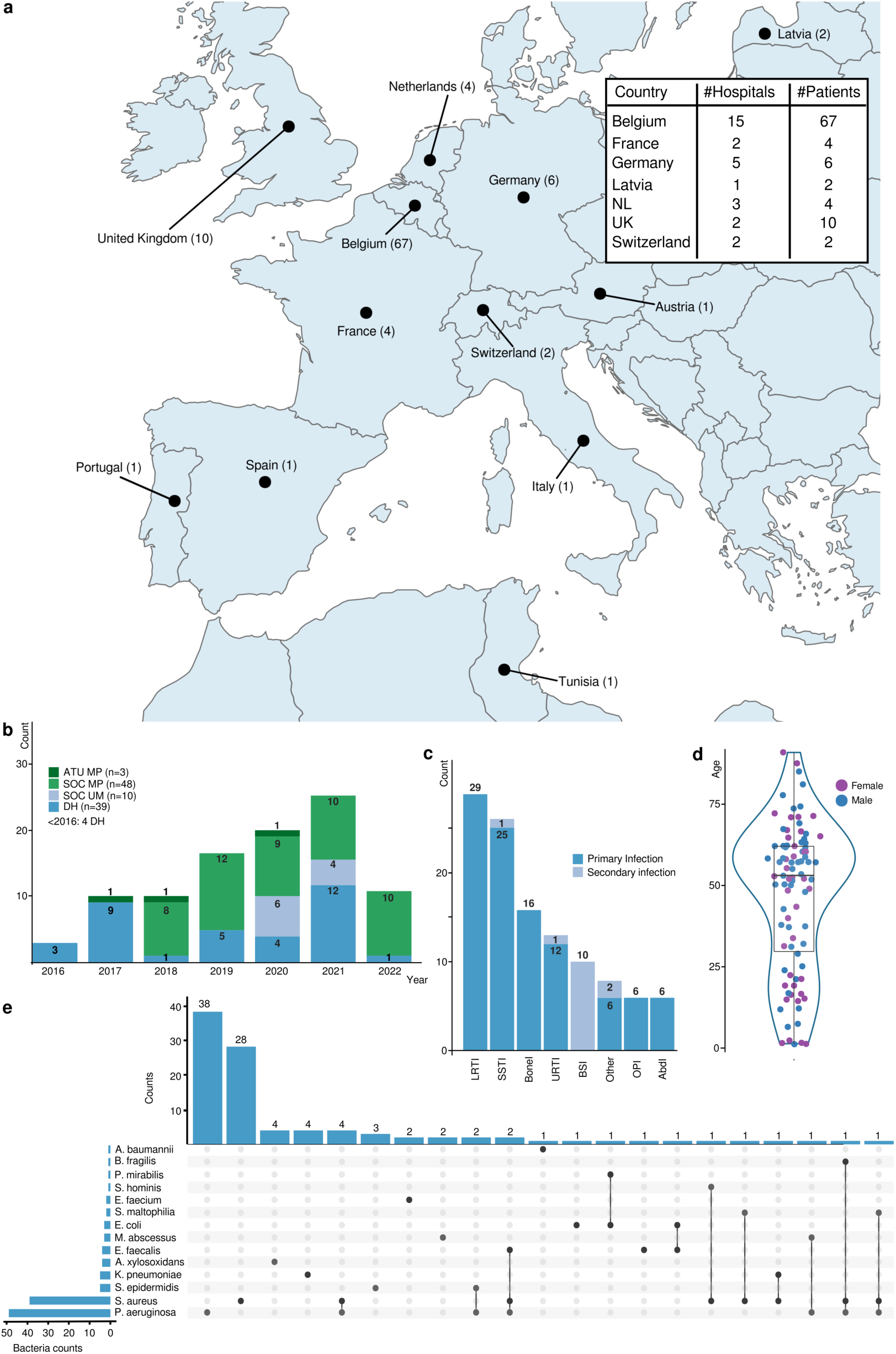
Characteristics of the patient population involved in the 100 consecutive bacteriophage therapy (BT) cases facilitated by the Belgian consortium. **a**, Geographic location of the BT cases. The table insert only concerns countries with more than one hospital where BT was performed. NL, The Netherlands; UK, United Kingdom. **b**, Number of BT cases, and their regulatory context, per year. SOC MP, standard-of-care with magistral bacteriophage preparations; DH, article 37 (unproven interventions in clinical practice) of the Declaration of Helsinki; SOC UM, standard-of-care with unlicensed medicines; ATU MP, “Autorisation Temporaire d’Utilisation” of magistral preparations. **c**, Primary and secondary (concomitant) infection types. AbdI, abdominal infection; BoneI, bone infection; BSI, bloodstream infection; LRTI, lower respiratory tract infection; OPI, orthopedic prostheses infection; SSTI, skin & soft tissue infection; URTI, upper respiratory tract infection. **d**, Patient age and gender distribution. Boxplot showing the interquartile range of the age (years) of the patients: first quartile (29.5), median (53), and third quartile (62). Female patients are represented by purple filled circles, male patients by blue filled circles. **e**, Targeted bacterial species. In some cases, bacteriophages targeted two or three bacterial species (connected by lines) in one patient.

### Bacteriophage preparations

One hundred and fourteen bacterial infections in 100 patients, caused by 14 bacterial species, were targeted by (combinations of) 26 individual bacteriophages (Extended Data Table 1) and six defined bacteriophage cocktails (Extended Data Table 2), including two commercially available cocktails (PyoPhage and IntestiPhage) produced by the Eliava Institute in Tbilisi (Republic of Georgia). Bacteriophages were provided by the QAMH and 13 Bacteriophage Donors** affiliated to eight institutes in six countries.

**Table 1.** General overview of bacteriophage therapy protocols according to the main infection types.

| Infection type | Application route | Bacteriophage carrier | Volume (mL) | Concentration (PFU/mL) | Dose | Duration |
| --- | --- | --- | --- | --- | --- | --- |
| Lower respiratory tract infections | Nebulization | NaCl 0.9% | 2–4 | $10^7$–$10^8$ | q6h | 5 days–6 weeks |
| Bone and orthopedic prostheses infections | Intralesional | NaCl 0.9% | 2–70 | $10^7$–$10^8$ | q24h | 5 days–3 weeks |
| Skin & soft tissue infections | Topical | NaCl 0.9% or Flaminal® Hydro | In excess | $10^7$–$10^9$ | q24h | 5 days–3 weeks |
| Upper respiratory tract infections | Nasal spray | NaCl 0.9% | 1–15 | $10^7$ | q8h | 1–3 weeks |
| Blood stream infections or other infection types* | Intravenous | NaCl 0.9% | 50–100 | $10^6$–$10^7$ | q24h | 5–10 days |

\*When the treating physician considered it was necessary to apply bacteriophages systemically.

**Table 2.** Results of the supportive tests performed for 21 of the present 100 consecutive bacteriophage therapy cases.

| Patient number | Infection type | Targeted bacterial species | Applied bacteriophage(s) | Bacteriophage administration route(s) | *In vivo* selection of bacteriophage resistance – possible underlying mechanism(s) | *In vitro* bacteriophage-antibiotic interactions | Bacteriophage immune neutralization | Clinical improvement | Eradication of targeted bacteria | Reference |
| --- | --- | --- | --- | --- | --- | --- | --- | --- | --- | --- |
| 9 | Fracture-related infection | *Klebsiella pneumoniae* | M1 | Intralesional (catheter) | Not observed | M1 synergy with ceftazidime/avibactam and meropenem | M1 neutralization emerged between days 8 and 18 after BT initiation | Yes | Yes | Eskenazi et al. (2022)<sup>8</sup> |
| 13 | Wound and bloodstream infection | *Pseudomonas aeruginosa* | 14-1, PNM and ISP (BFC 1) | Topical and intravenous | Not observed | No concomitant antibiotics | 14-1 neutralization emerged 10 days after BT initiation* | Yes | Yes | Jennes et al. (2017)<sup>25</sup> |
| 16 | Cystic fibrosis lung transplant infection | *Achromobacter xylosoxidans* | JWAlpha, JWDelta, JWT and 2-1 (APC 1.1 and APC 2.1) | Nebulization | Yes – p.Tyr601X MS Mut in colicin I receptor Cir | No concomitant antibiotics | NSA | Yes | Yes | Lebeaux et al. (2021)<sup>26</sup> |
| 20 | Liver transplant and bloodstream infection | *P. aeruginosa* | 14-1, PNM and ISP (BFC 1) | Intralesional (infusions) and intravenous | Yes – p.Asp388Ala MS Mut in PilB and deactivation of PilM by insertion of IS5 transposase, both involved in Type IV pili biosynthesis, without impact on virulence | PNM synergy with colistin, aztreonam, and gentamycin | ISP neutralization emerged five weeks after BT initiation. No neutralization of 14-1 or PNM | Yes | Yes | Van Nieuwenhuijse et al. (2022)<sup>27</sup> |
| 21 | Bone allograft infection | *Staphylococcus aureus* | 14-1, PNM and ISP (BFC 1) | Intralesional (catheter) and intravenous | Not observed | ISP synergy with clindamycin, additive effect of ISP and ciprofloxacin, moderate ISP antagonism with rifampicin | NSA | Yes | Yes | Van Nieuwenhuijse et al. (2021)<sup>28</sup> |
| 22–24 and 42 | Orthopedic infections | *P. aeruginosa*, *S. aureus*, *S. epidermidis*, and *Enterococcus faecalis* | 14-1, PNM, ISP (BFC 1) and PyoPhage | Intralesional (catheter) | NSA | NA | One month after BT initiation, no bacteriophage neutralization could be detected | Yes | Yes | Onsea et al. (2019)<sup>29</sup> |
| 26 | Spinal infection | *P. aeruginosa* | 4029, 4032, and 4034 | Local and intravenous | Not observed | NA | NSA | Yes | Yes | Ferry et al. (2022)<sup>30</sup> |
| 27 | Orthopedic infection | *P. aeruginosa* | 14-1, PNM, and ISP (BFC 1) | Local | Not observed | Additive effect of the bacteriophage cocktail with ceftazidime/avibactam | NSA | Yes | Yes | Racenis et al. (2022)<sup>31</sup> |
| 30 | Chronic sinusitis | *P. aeruginosa* and *S. aureus* | 14-1, PNM and ISP (BFC 1) | Nasal spray | Yes – p.Ala154Pro MS Mut in PilC, involved in Type IV pili biosynthesis | No concomitant antibiotics | NSA | No | No | Unpublished |
| 43 | Liver transplant infection | *Enterococcus faecium* | EFgrKN and EFgrNG | Intravenous | Not observed | Synergy of EFgrKN with vancomycin, loss of vancomycin resistance | Forty-nine days after BT initiation, no bacteriophage neutralization could be detected | Yes | No | Paul et al. (2021)<sup>33</sup> |
| 54 | Ventilator associated pneumonia | *P. aeruginosa* | 14-1, PNM and PT07 | Nebulization | Yes – p.Thr230Proline MS Mut in PilR, involved in Type IV pili biosynthesis | No clear interaction between PNM or 14-1 and colistin | Two months after BT initiation, no bacteriophage neutralization could be detected | Yes | No | Unpublished |
| 55 | Musculoskeletal infection | *S. epidermidis* | ISP and BE06 | Intralesional and intravenous | Not observed | No concomitant antibiotics | NSA | No | No | Unpublished |
| 64 | Anal fistula | *P. aeruginosa* | 14-1, PNM and PT07 | Intralesional | Yes – Selection of another strain, which is not a host for bacteriophages 14-1, PNM, or PT07 | No concomitant antibiotics | Four and seven months after BT initiation, no bacteriophage neutralization could be detected | Yes | Yes | Unpublished |
| 66 | Cystic fibrosis lung infection | *M. abscessus* | 8UZL | Nebulization and intravenous | NSA | NA | 8UZL neutralization emerged seven days after BT initiation | No | No | Unpublished |
| 71 | Lung infection | *P. aeruginosa* | PT07 | Nebulization | Not observed | Additive effect of PT07 and ceftazidime | NSA | Yes | Yes | Unpublished |
| 82 | Cystic fibrosis lung infection | *P. aeruginosa* | 4P and DP1 | Nebulization | Yes – Selection of another strain, which is not a host for bacteriophages 4P and DP1 | Synergy of 4P and DP1 with levofloxacin; no clear interaction between 4P or DP1 and tobramycin | NSA | Yes | Yes | Unpublished |
| 91 | Lung infection | *P. aeruginosa* | 14-1, PNM and PT07 | Nebulization and intravenous | Yes –<br>**LPS biosynthesis:**<br>Is 2 and 3) p.Trp139X NS Mut in WapH, Is 2 and 3) p.Gln239X NS Mut in GalU, Is 4 and 5) p.Leu162Pro MS Mut in WapR, Is 4 and 5) p.Leu60_Leu63del in WbpR<br>**Type IV pili biosynthesis:**<br>Is 4 and 5) p.Arg120fsX in FimV, missing the first 165 AA<br>**Other:**<br>Is 6) p.Gly406Ser MS Mut in CupE5 fimbriae assembly protein, Is 2, 3, 4, 5, and 6) p.Arg994Gly MS Mut in MexB of MexAB-OprM, Is 4 and 5) p.His87Asp MS Mut in GyrA | PT07 synergy with colistin and meropenem | Two weeks after BT initiation, no bacteriophage neutralization could be detected | Yes | No | Unpublished |
| 92 | Generalized necrotizing fasciitis, empyema, bacteremia | *S. aureus*, *P. aeruginosa*, and *Stenotrophomonas maltophilia* | ISP, 14-1, PNM, PT07 and BUCT700 | ISP: Intravenous, Intrapleural, intraperitoneal, and nebulization; All: topical | Not observed | ISP synergy with vancomycin, ceftarolin and clindamycin | ISP neutralization emerged 6 days after BT initiation | Yes | Yes | Unpublished |

AA, amino acids; BT, bacteriophage therapy; del, deletion; fs, frameshift; Is, isolate; MS Mut, missense mutation; NA, not analyzed; NS Mut, nonsense mutation; NSA, no samples available; X, stop.

#### Quality and safety

Sciensano controlled the quality and safety of 43 batches of individual bacteriophage active pharmaceutical ingredients (APIs) produced by the QAMH. These batches exhibited an average bacteriophage titer of 8.34 x 10^9^ plaque forming units (PFU)/mL (standard deviation (SD) 1.16 x 10^10^), a pH of 7.32 (SD 0.037), a bioburden of 0 colony forming units (CFU)/mL (SD 0) and a median endotoxin level of 5 endotoxin units (EU)/mL (SD 89.14). The endotoxin limit for the bacteriophage preparations was defined on the basis of dosage and on the patient’s weight. The administered endotoxin doses were, irrespective of the administration route, always well below the threshold pyrogenic dose for intravenous administration, i.e. < 5.0 EU of endotoxin per kilogram of body mass per hour. Bacteriophage genomes contained no genetic determinants known to confer lysogeny, toxicity, virulence, or antibiotic resistance. Host bacteria used in the manufacturing process were as safe (or least pathogenic) as possible. Some production hosts were shown to contain prophages. Bacteriophage productions with > 5% of sequencing reads derived from actively replicating prophages were not used in therapy.

#### Pre-adaptation of bacteriophages

The most frequently used bacteriophages, i.e. *Staphylococcus* bacteriophage ISP (33 patients), and *P. aeruginosa* bacteriophages 14-1 (22 patients), PNM (21 patients), and PT07 (18 patients) (Extended Data Table 1), were regularly (one to two times per year) adapted using a selection of three to five recent bacterial strains of concern. In addition, 13 bacteriophages were specifically pre-adapted to lyse the patient’s bacteria in a therapeutically relevant manner (Methods) – that is, to produce stable lysis (without emergence of bacteriophage-insensitive mutants) in liquid culture, for typically 48 hours, at a multiplicity of infection (MOI) ≤ 1 (Extended Data Table 3). We considered the relative efficiency of plating (EOP) as a relative measure of lysis efficiency, which – in this context – is defined as the lytic activity (titer) of the bacteriophage on the patient’s bacterial strain, divided by the titer observed in a reference bacterial host known to be highly susceptible to the bacteriophage. A therapeutically acceptable bacteriophage should have an EOP ≥ 0.1 on the patient’s bacterial strain. The genomes of the pre-adapted bacteriophages are sequenced and analyzed and compared to those of their original precursors, as part of Sciensano’s SAPHETY project (https://www.sciensano.be/en/control-and-safety-assessment/safety-therapeutic-bacteriophage-preparations), which focuses on setting new standards for the quality and safety of therapeutic bacteriophage products. A detailed genetic comparison of all patient-adapted phages falls outside the scope of this study. However, two sequence-based comparison between the pre-adapted and the original variant of the bacteriophage were previously reported. The comparison between *Klebsiella pneumoniae* bacteriophage M1 and its pre-adapted version was recently published^8^. It was suggested that a missense mutation in the loop region of the hinge connector of the distal tail fiber protein might have caused alterations in the bacteriophage receptor, leading to a better interaction with the bacterial receptor and improved lytic activity against the patient’s *K. pneumoniae* isolates. Another pre-adaptation effort concerned increasing the activity of *S. aureus* bacteriophage ISP against a *S. epidermidis* clinical isolate in view of personalized BT. The pre-adaptation process (four serial passages) resulted in missense mutations in genes predicted to code for a receptor binding protein, a carbohydrate-binding domain protein, an uracil-DNA glycosylase, and a hypothetical protein (Extended Data Fig. 2). Staphylococcal Twort-like bacteriophages like ISP are known to harbor at least two receptor binding proteins, binding both the glycosylated wall teichoic acids (WTAs) and the WTA backbone^37^. It is likely that the first two missense mutations, which occurred in genes that were most closely related (closest BLAST hits) to those coding for these WTA and glycosylated WTA binding proteins, enabled ISP to better recognize the specific ISP receptor variants on the cell wall of the patient’s *S. epidermidis* isolates. However, the increased virulence and resistance suppression of the pre-adapted ISP variant was accompanied by a decreased host range. Where the original ISP clone showed a moderate activity against 3/16 *S. epidermidis* strains, the adapted variant showed a therapeutically acceptable activity against the patient’s strain only. The host range of Sb-1, another *S. aureus* Twort-like bacteriophage, was expanded by adaptation to previously resistant clinical isolates. Comparative genomic analysis between the parental Sb-1 bacteriophage and two expanded host range mutants revealed a hypervariable complex repeat structure in the Sb-1 genome with a distinct allele correlating with the host range expansion^38^.

### Bacteriophage therapy protocols

Most Bacteriophage Therapy Providers* by and large adhered to the BT protocols that were proposed by the physicians of the QAMH, and were based on the application instructions endorsed by the Ministries of Health and of Medical and Microbiology Industry of the USSR^21–23^, and on the long-time (primarily empirical) experience of the Eliava Institute. This resulted in a surprisingly small variation in BT protocols within a given indication. Table 1 provides a general overview of these protocols, while the individual protocols of the 100 cases are listed in Supplementary Table 1.

### Supporting assays

For 21 patients, bacterial samples and/or serum samples were provided, allowing assessment of (i) the potential in vivo emergence of resistance against the applied bacteriophages, ii) in vitro bacteriophage-antibiotic interactions, and/or iii) the emergence of bacteriophage immune neutralization (Table 2).

#### Selection of bacteriophage resistance

We observed the in vivo selection of bacterial strains exhibiting a bacteriophage-insensitive phenotype, and the possible underlying phenotype-genotype associations, in seven of 16 (43.8%) patients for which adequate follow up bacterial samples were available for testing (patients 16, 20, 30, 54, 64, 82, and 91 in Table 2). Whole genome single nucleotide polymorphism (SNP) analysis was performed for bacterial isolates from the patients where bacteriophage insensitivity emerged. In two patients (64 and 82 in Table 2), sequential bacteriophage-susceptible and bacteriophage-insensitive *P. aeruginosa* isolates were determined not to be clonal. Phylogenetic comparison showed that for patient 82, bacteriophage-susceptible strains belonged to an emerging rare sequence type (ST)235, whereas bacteriophage-resistant strains belonged to the more prevalent multidrug-resistant ST357 (Table 2 and Fig. 2a)^39,40^. For patient 64, the susceptible strain was ST1233 (same ST as the strains from patient 91), while the resistant strain was determined to be ST549 (Table 2 and Fig 2a). In these two patients, BT likely selected for *P. aeruginosa* strains that were not a suitable host for the applied bacteriophages. Clinical improvement was reported in both patients.

**Fig. 2.**
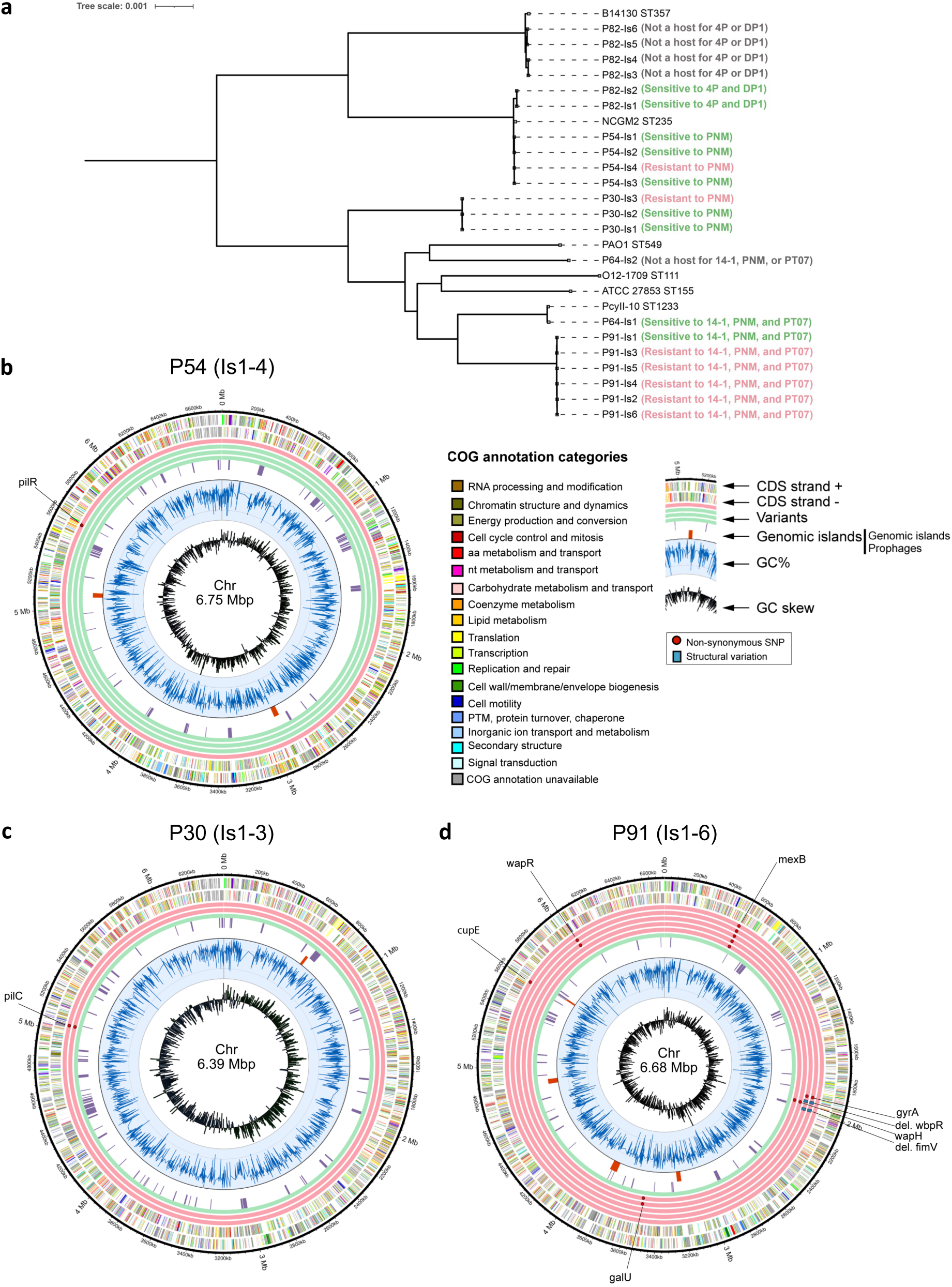
The *in vivo* emergence of bacteriophage resistance during bacteriophage therapy (BT), as monitored by whole genome analysis of sequential bacterial isolates, in patients 30, 54, 64, 82, and 91 (in vivo emergence of bacteriophage resistance in patients 16 and 20 was previously discussed elsewhere (Table 2)). **a**, Maximum likelihood phylogenetic tree of the genomes of the analyzed sequential bacterial isolates. **b**, Circular chromosomic view (CCV) of the bacterial genomes of sequential isolates (Is) of *Pseudomonas aeruginosa* strains retrieved just before (Is1, inner circle) and during BT (Is2-6) from patients 30, 54, and 91. Green rings display the genomes of bacteriophage-susceptible isolates, while the red rings display the genomes and relevant (for bacterial bacteriophage resistance) mutations in bacteriophage-resistant isolates. The two multi-colored outer rings display the protein annotations (categories) as present in the database of Clusters of Orthologous Groups of proteins (COGs). aa, amino acid; bp, basepairs; CDS, coding sequence; IS, insertion sequence; Mb, megabases; nt, nucleotide; PTM, post-translational modification; SNP, single-nucleotide polymorphism.

SNPs or deletions in genes related to the bacteriophage receptor were assumed to be at the basis of the resistance phenotype in five patients (16, 20, 30, 54, and 91 in Table 2). In three of them (patients 30, 54, and 91), the targeted *P. aeruginosa* strains were not eradicated. The selection of bacteriophage-resistant mutants in two patients (16 and 20) was previously described^26,27^. In patient 16, an isolate of the targeted *Achromobacter xylosoxidans* strain emerged to harbor a missense mutation in the gene coding for the colicin I receptor Cir, which was identified as a bacteriophage receptor. In patient 20, a missense mutation occurred in the *pilB* gene of the targeted *P. aeruginosa* strain, while the *pilM* gene was inactivated by the insertion of IS5 transposase. Both *pilB* and *pilM* are involved in the biosynthesis of Type IV pilus (T4P), the receptor for the applied *P. aeruginosa* bacteriophage PNM.

Bacteriophage-resistant *P. aeruginosa* mutants were also isolated from patient cases 30, 54 and 91 (Table 2 and Fig 2b-d). Among these mutations, SNPs were identified that corresponded to regions related to T4P in all three patients. In one patient (54), this mutation was in the *pilR* gene, coding for the transcriptional activator of a two-component system that regulates expression of the major pilin subunit PilA (Fig. 2b)^41^. In another patient (30) isolate this mutation was in a gene coding for an inner membrane component, PilC, essential for T4P biogenesis (Fig. 2c)^42^. For patient 91, a premature stop codon was introduced producing a truncated gene variant of the gene *fimV*, which expresses a part of the inner membrane assembly of T4P in *P. aeruginosa* (Fig. 2d)^43^. In addition, this patient was shown to harbor bacteria that exhibited simultaneous resistance to all three unique *P. aeruginosa* bacteriophages from treatment, PNM, PT07, and 14-1 (Table 2 and Fig. 2d). Interestingly, we observed two distinct bacteriophage-resistant variants of the initially targeted *P. aeruginosa* strain, each showing resistance to the three applied bacteriophages. Surprisingly, the three bacteriophages had different bacterial receptors. *P. aeruginosa* bacteriophage 14-1 infects via a lipopolysaccharide (LPS) receptor^44^. Not surprisingly, SNPs were identified in genes in the outer core of the *P. aeruginosa* LPS membrane, i.e. *wapH*, *galU*, *wbpR* (gene products truncated in these three mutant variants) and *wapR*. Although the receptor for *P. aeruginosa* bacteriophage PT07 is not known, sequence similarity to PAK-P1 like bacteriophages (98.26% identity to bacteriophage PaP1) suggests this bacteriophage is dependent on the *P. aeruginosa* MexAB-OprM multidrug efflux pump^45^. A resistance mutant of PT07 was identified with a SNP in the gene *mexB*. Two *P. aeruginosa* isolates (Is 4 and 5 in Table 2) from patient 91 had both the *mexB* mutation and another mutation in DNA gyrase subunit A (*gyrA*), part of the bacterial DNA topoisomerase. This mutation (H87A) is within the GyrA quinolone-resistance determining region (QRDR)^46–48^. The interplay of the MexAB-OprM efflux pump and a DNA gyrase mutation has been associated with high-level fluoroquinolone resistance in *P. aeruginosa*^49^. Interestingly, the bacteriophage-insensitive *P. aeruginosa* isolates retrieved from patient 91 carrying the double mutation in *mexB* and *gyrA* showed a re-sensitization to fluoroquinolones, illustrated by a decrease in minimum inhibitory concentration (MIC) form ≥ 4 to 0.5 µg/mL for ciprofloxacin, and from ≥ 8 to 1 µg/mL for levofloxacin. Of note, patient 91 was treated concomitantly with bacteriophages and the antibiotics meropenem, colimycin and vancomycin.

*Galleria mellonella* larvae were used as an infection model to readily assess the virulence of bacteriophage-susceptible versus bacteriophage-resistant variants of *P. aeruginosa* strains isolated from patients 30, 54 and 91. Larvae infected with original, bacteriophage-susceptible, isolates showed rapid and significant mortality within 48 hours (100% death) (Extended Data Fig. 3). The groups infected with bacteriophage-resistant mutants from patient 91 with multiple mutations (> 2) in genes encoding for different regions (LPS, MexAB-OprM, and/or T4P and/or DNA gyrase) showed significantly higher survival rates (*P* = 0.004 and *P* < 0.0001) compared to the larvae infected with the original isolate in this model system. After 24 hours, significantly higher survival rates were also observed for the larvae infected with the bacteriophage-resistant variant of the *P. aeruginosa* strain isolated from patient 30 as compared to the original bacteriophage-susceptible variant (*P* = 0.01). However, all larvae from these two groups died in 48 hours. The larvae infected with the bacteriophage-resistant isolate from patient 54 showed no difference in survival compared to ones infected with the original isolate.

#### In vitro bacteriophage-antibiotic interactions

In vitro bacteriophage-antibiotic interaction experiments revealed a synergistic or additive effect of bacteriophages and the concomitantly applied antibiotics in nine out of ten evaluated patients (9, 20, 21, 27, 43, 71, 82, 91, and 92). An overview of the test results is presented in Table 2. The results of the experiments concerning the first five patients (9, 20, 21, 27, and 43) were reported previously^8,27,28,31,33^. The detailed results (OmniLog^®^ growth curves) for the five most recent patients (54, 71, 82, 91, and 92) are presented in Fig. 3. In vitro synergy with bacteriophages was observed for nine antibiotics (aztreonam (patient 20), ceftarolin (92), ceftazidime/avibactam (9), clindamycin (21 and 92), colistin (20 and 91), gentamicin (20), levofloxacin (82), meropenem (9 and 91), and vancomycin (43 and 92), and an additive effect for three antibiotics (ceftazidime/avibactam (27), ceftazidime (71), and ciprofloxacin (21). For one patient (54), no significant in vitro interactions between colistin and *P. aeruginosa* bacteriophages PNM (Fig. 3a) or 14-1 (Fig. 3b) were observed. Bacteriophages 4P and DP1 acted in synergy with levofloxacin (Fig. 3d-e), but showed no clear interaction with tobramycin (Fig. 3f-g), when tested in vitro against the *P. aeruginosa* strain of patient 82. Of note, a moderate antagonism was observed for *S. aureus* bacteriophage ISP with rifampicin (patient 21 in Table 2) in one of our previously published BT cases^28^.

**Fig. 3.**
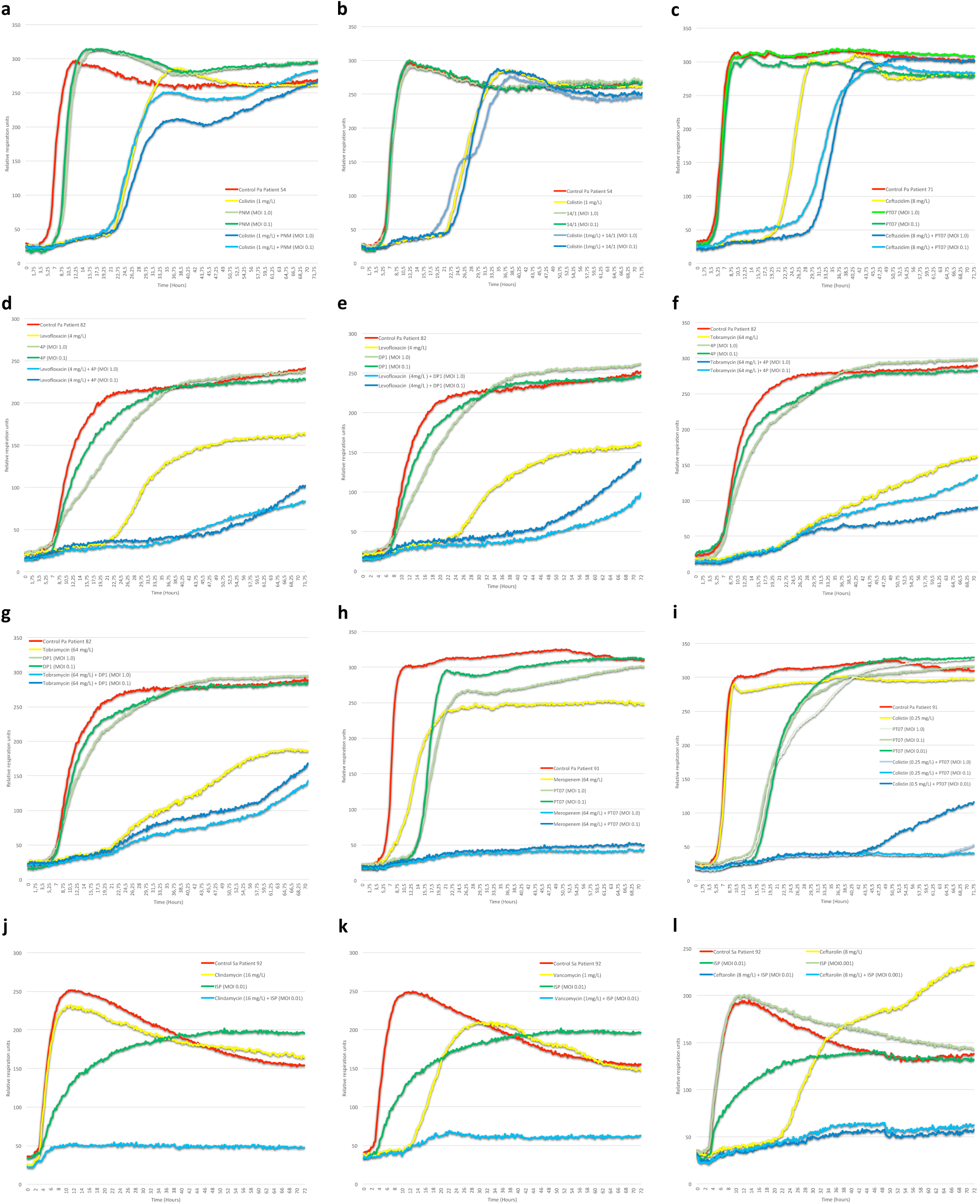
Results of the in vitro evaluation of the combined effects of bacteriophages and concomitantly applied antibiotics on the targeted bacterial strains, as determined by an OmniLog^®^ system, for patients 54, 71, 82, 91, and 92 (those for patients 9, 20, 21, 27, and 43 were previously discussed elsewhere (Table 2)). Bacterial proliferation is presented through relative units of cellular respiration. **a-b**, No additive effect of colistin and bacteriophages PNM (**a**) and 14-1 (**b**) for patient 54. **c**, Additive effect (delayed bacterial growth) of ceftazidime and bacteriophage PT07 for patient 71. **d-e**, Synergistic effect of levofloxacin and bacteriophages 4P for patient 82 (**d**) and DP1 (**e**). **f-g**, No additive effect of tobramycin and bacteriophages 4P (**f**) and DP1 (**g**) for patient 82. **h-i**, Synergistic effect of bacteriophage PT07 and the antibiotics meropenem (**h**) and colistin

#### Bacteriophage immune neutralization

Bacteriophage immune neutralization was observed, between six and 35 days after initiation of BT, in five of 13 (38.5%) screened patients (9, 13, 20, 66, and 92 in Table 2 and Fig. 4a-d). Bacteriophage immune neutralization always involved invasive (intravenous and/or intralesional) bacteriophage administrations. In four of these five cases (patients 9, 13, 20 and 92), clinical improvement and eradication of the targeted bacterial pathogen were nevertheless observed. In a liver transplant patient (43 in Table 2), the intravenous administration of bacteriophages did not elicit any immune neutralization. In another liver transplant patient (20), bacteriophage immune neutralization emerged, but only after five weeks, and it concerned only one of the three bacteriophages that had been applied (Table 2 and Fig. 4c).

**Fig. 4.**
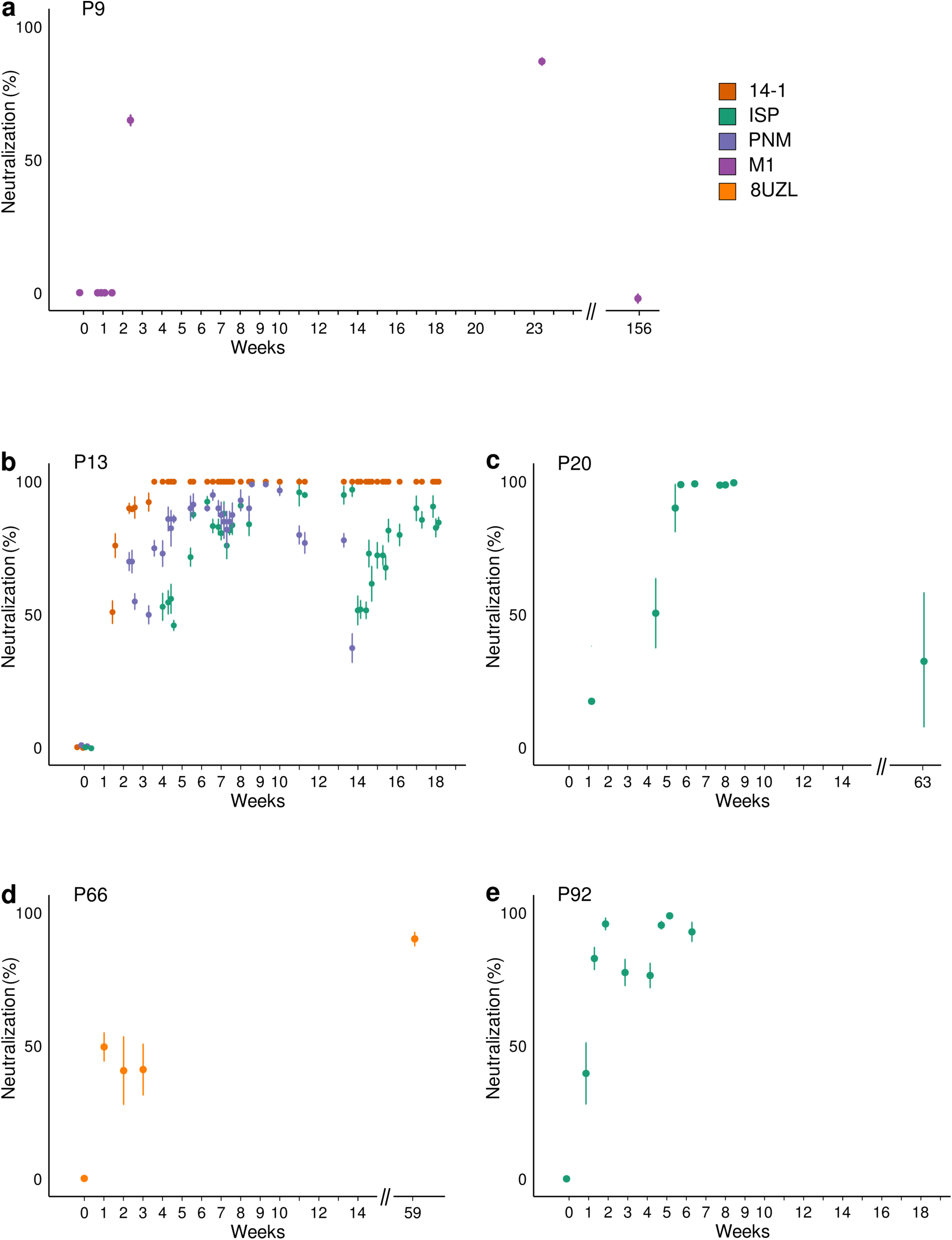
Emergence of bacteriophage immune neutralization. Chronological bacteriophage immune neutralization (BIN) activity against the applied bacteriophages in sera collected before, during and after bacteriophage therapy (BT) in patients 9 (**a**), 13 (**b**), 20 (**c**), 66 (**d**), and 92 (**e**). The evolution over time of the serum BIN activity against the applied bacteriophages is shown as % bacteriophage titer loss (compared to pre-BT control sera) after incubation of the bacteriophages with sequential serum samples for 30 min. BIN activity appeared 1 to 5 weeks after BT initiation. Data are presented as mean values of three biological replicates with error bars representing the standard deviation of the means.

### Clinical outcome

#### Clinical improvement

Clinical improvement was reported for 77.2% (88/114) of targeted infections and eradication of the targeted bacteria was observed in 61.3% (65/106) of infections for which relevant bacteriological follow up data was available (Supplementary Table 1). For eight targeted infections, in eight patients, no adequate post BT bacteriological data was available (Supplementary Table 1). BT resulted in clinical improvement without bacterial eradication in 18 (17.0%) of 106 targeted and bacteriologically monitored infections (Supplementary Table 1). Conversely, in two patients (44 and 93 in Supplementary Table 1), eradication of the targeted pathogens was observed without clinical improvement. In patient 44, an infection with an additional (non-BT-targeted) bacterial species (*Acinetobacter baumannii*) surfaced during BT, which ultimately resulted in an *A. baumannii* pulmonary septic shock and the patient’s death, and this despite intravenous administration of tigecycline. Patient 93 succumbed to tumor progression.

For 21 (22.8%) of the 92 patients for which bacteriological follow up data was available, no clinical improvement nor bacterial eradication could be observed. Five of these patients (3, 36, 40, 69, and 96 in Supplementary Table 1) died due to septic shock (*n* = 2), cardiogenic shock (*n* = 1), multi-organ failure (*n* = 1), or COVID-19 infection (*n* = 1). In 69.3% (79/114) of targeted infections, concomitant standard-of-care antibiotics were administered (Supplementary Table 1).

Fisher exact test for count data showed univariate significant effects on eradication for the following categorical variables: concomitant use of antibiotics, antibiotic resistance profile of the targeted bacteria and the clinical setting. No effects of age or gender on eradication were found using a logistic regression analysis. A stepwise logistic regression analysis of eradication on all independent variables, determined that the concomitant use of antibiotics (variable ABCONCOM) was the most informative variable in the reduced dataset (Extended Data Fig. 4 and Supplementary Table 2). Eradication was 70% less probable when no concomitant antibiotics were used (odds-ratio = 0.3; 95% confidence interval = 0.127–0.749). A sketch (Extended Data Fig. 4.a) and a confusion matrix (Extended Data Fig. 4.b) show that our logistic regression model is right 65% (40+20/92) of the time. The antibiotic resistance profile of the target bacteria (ABRPROF) and the clinical setting (CLINSETT) as well as their interactions with the concomitant use of antibiotics (ABCONCOM) were not significant in the overall logistic regression model. This is attributable to confounding relations between these three variables within this dataset. A highly significant association was found between clinical improvement and bacterial eradication. Eradication is 96% more probable after clinical improvement. Of the 23 patients with no clinical improvement, only two patients expressed eradication. Of the 69 patients with clinical improvement, 53 had full eradication. Intravenous BT, as stand-alone or concomitant therapy, was not shown to significantly impact clinical outcome, nor did patient age or gender, the persistence of the bacterial infection (chronic or acute), or the use of more than one targeting bacteriophage per bacterial strain. Clinical improvement or bacterial eradication were not significantly correlated with the presence of either *P. aeruginosa* or *S. aureus*, where other species were not considered separately, as their prevalence in this study population was too low, or with any individual bacteriophage or bacteriophage cocktail. Obviously, considering the relatively high number of combined categorical and numerical variables in the analyzed data, the majority of patients were unique cases in most of the variables. As a result, on this dataset, no inferential statistics could be applied, because these data were not a random, nor a representative sample of a population of BT-treated patients. Any data analysis can as such only be interpreted as information pertaining to the analyzed patient population.

#### Adverse events

Fifteen adverse events were reported, including seven mild to moderate adverse reactions possibly linked to BT (Extended Data Table 4). All adverse reactions resolved. No correlation between adverse events and a bacteriophage product or application route could be made.

## Discussion

Facing an impending antibiotic resistance crisis, a BT renaissance in the Western world was announced about two decades ago^4^. Unfortunately, since then, no bacteriophage medicines have made it – through the conventional drug development funnel – to the market. In addition, most bacteriophage preparations that are currently developed by pharmaceutical companies are defined bacteriophage cocktails, which underappreciate a number of peculiarities of bacteriophages, including their target specificity and antagonistic coevolution with their bacterial hosts^6^. In recent RCTs, these invariable, defined bacteriophage cocktails showed disappointing results, which contrast with those of an increasing number of case studies using selected, or even pre-adapted bacteriophages as adjunctive therapy^6^.

From the moment we developed promising therapeutic bacteriophage preparations, in 2007, we faced one of the fundamental ethical dilemmas of medical research: should we pursue scientific precision through controlled clinical trials, or should we immediately provide bacteriophages to patients in dire need? Taking into account the numerous empirical indications that BT can be efficacious^50^ and some ethical considerations^51^, and mindful of the natural tendency of medical staff to help patients, we opted to do both. Therefore, in Belgium, a comprehensive BT consortium was formed to meet, to some extent, the growing demand for BT, and to perform prospective clinical trials^20^. In addition, in 2018, a dedicated BT regulatory framework was put in place in Belgium^19^. The Belgian consortium decided not to re-invent BT, but rather to build on the one-century-long – to some extent empirical – experience of the Eliava Institute.

In this overview of the first 100 consecutive cases facilitated by this consortium, we show that: (i) we were able to produce more than forty batches of personalized bacteriophage APIs, which were subsequently certified for use in pharmaceutical preparations; (ii) when used in the treatment of 114 difficult-to-treat infections of various types and etiology, in combination with antibiotics in 69.3% of cases, these preparations lead to clinical improvement in 77.2%, and eradication of the targeted bacteria in 61.3% of cases; (iii) seven mild to moderate adverse reactions were reported.

The overwhelming representation of *P. aeruginosa* (44% of patients) and *S. aureus* (35% of patients) as targeted bacterial species is because these are overall major causes of severe nosocomial infections, but also the main microorganisms causing invasive burn wound infection^52^, which is historically a major focus of attention of the infectiologists of the QAMH, where the first bacteriophage treatments were carried out^53^.

The treatment of 100 patients required combinations of 26 individual bacteriophages, of which 13 were pre-adapted to more efficiently target the patients’ infections. Importantly, we adhere to the Eliava Institute’s standards for what constitutes an efficient therapeutic bacteriophage, namely one that is capable of stable lysis (without emergence of bacteriophage resistance) of the targeted bacterial strain(s) for a prolonged period of time (typically 24–48 hours) and at a low MOI (typically ≤ 1). The additional 40 patients treated (clinical follow up in progress) after the 100 patients in this formal retrospective analysis, required six additional bacteriophages. Pre-adapting bacteriophages to a bacterial pathogen was shown, in vitro, to increase pathogen clearance and to lower resistance evolution in *P. aeruginosa*^54^. It is often forgotten that the BT pioneers used bacteriophages specifically directed against the patients’ infecting bacteria. In 1921, the Belgian physician René Appelmans published a method (the “Appelmans protocol”), which since then was used for the directed in vitro evolution of therapeutic bacteriophages in institutions where traditional BT was practiced, such as the Eliava Institute^55,56^.

Of note, all bacteriophage preparations were offered free of charge, with most R&D and manufacturing costs borne by the Royal Higher Institute for Defense, the QAMH, Sciensano, and KU Leuven. However, this endeavor – providing 43 batches of 26 bacteriophages for the treatment of 100 patients – would not have been possible if one had to comply with the large body of costly and time-consuming requirements and procedures for GMP manufacturing and licensing of biological medicinal products. Companies focusing on defined bacteriophage preparations, developing a few “easy” bacterial species (from a bacteriophage promiscuity perspective) for use in commercially viable indications (e.g. concerning a large number of patients), might be able to deal with the demanding requirements of the conventional medicinal product (drug) licensing pathway, including GMP-certification, pre-clinical test, and clinical trials. However, for a BT center wanting to help all patients in need, irrespective of the concerned indications or bacterial species, these requirements form an insurmountable barrier in terms of timelines and cost.

We experienced first-hand how more elaborate and logistically complex personalized BT concepts are, compared to one-size-fits-all approaches, with bacterial strains and matching bacteriophages being exchanged between dozens of institutes in 12 countries. This experienced disadvantage of the personalized BT approach is partly responsible for the current focus of our R&D on the development of an instant and on-site production system for bacteriophages, based on artificial intelligence and synthetic biology approaches^57^. The methods currently used to produce therapeutic bacteriophage preparations have barely evolved since 1919, when Félix d’Hérelle exploited for the first time the therapeutic potential of phages, while biology has evolved to provide technologies such as artificial intelligence (AI) and synthetic biology, which might solve phage specificity and bacterial phage resistance issues.

BT protocols were based on the extensive expertise present in the former USSR^21–23^, and more specifically in the Eliava Institute. These protocols prescribe relatively low bacteriophage doses, usually around 10^7^ PFU/mL, and ranging from10^6^–10^7^ PFU/mL for continuous intravenous BT to 10^9^ PFU/mL for topical BT in a few SSTI cases. In contrast, some clinics prefer the administration of considerably higher doses, for instance up to 10^10^–10^11^ PFU/mL for intravenous BT^12,58^. In the US, the Antibacterial Resistance Leadership Group (ARLG) Phage Taskforce suggests using the highest safe and tolerated dose of a bacteriophage product with endotoxin levels below the acceptable limits set by the FDA, to maximize bacteriophage concentrations at the site of infection and infect as many host cells as possible with the first dose^59^. The ARLG Phage Taskforce, however, acknowledges that clinical outcomes are not always improved with higher doses, reflecting the complexity of effective bacteriophage dosing. We observed an increase in in vitro bacteriophage efficiency (lytic activity) with increasing MOI, up to a certain MOI, after which regrowth can be observed more frequently and at an earlier point in time (Extended Data Fig. 5). The effective bacteriophage doses in the body are also determined by the route of bacteriophage administration. Most established BT protocols presented here are based on the principle that bacteriophages are best administered directly into the site of infection. Oral administrations were not used because no gastrointestinal infections were treated.

In 17% (18/106) of targeted infections, for which bacteriological follow up data was available, clinical improvement was reported even though the targeted bacteria were not eradicated. This phenomenon was also observed at the Eliava Institute (Chanishvili N, unpublished) and may be linked to a reduced virulence of the selected bacteriophage-resistant bacteria.

Infecting bacteriophages are confronted with a plethora of anti-bacteriophage defense mechanisms^60^. In addition, already in 1943, Salvador Luria and colleagues reported on the emergence of bacteriophage-resistant bacterial mutants in liquid cultures^61^. They observed that when a bacterial culture is attacked by a suitable lytic bacteriophage, the culture will clear, but after further incubation for a few hours to sometimes days, the culture can become turbid again, due to the growth of a bacterial variant, which is resistant to the applied bacteriophage.

The *in vivo* emergence of bacteriophage resistance is perceived as a major issue for BT, and this while, apart from a few sporadic reports^7,27^, little is known about its prevalence, and potential persistence and spread in the (hospital) environment. In *P. aeruginosa* endocarditis models, bacteriophage resistant mutants emerged in vitro (fibrin clots), but not in vivo (rats). The in vitro selected resistance mutations occurred in bacterial surface determinants important for infectivity^62^. There are, however, several reasons to anticipate that bacteriophage resistance could be different in vivo. The human body is bound to be a more stressful environment for bacteria due to limited resources (e.g. nutrients), the presence of an immune system, and competing microbes. Recently, Castledine and colleagues observed a parallel evolution of bacteriophage resistance and of virulence loss in *P. aeruginosa* response to bacteriophage treatment (in one patient), and this in vivo as well as in vitro^63^. In vivo selected resistance was associated with reduced growth rates, whereas in vitro selected bacteriophage-resistant isolates evolved greater biofilm production. They also suggested that resistance resulted from selection of de novo mutations rather than sorting of existing variants. Westra et al. showed that when bacteriophage infection risk is high, constitutive resistance mechanisms, such as a mutation of the bacteriophage receptor, are selected by the bacterial hosts, rather than inducible resistance mechanisms, such as a CRISPR (clustered regularly interspaced short palindromic repeats) system^64^. They suggested that the reason for this is that constitutive mechanisms come with a fixed cost that does not alter, irrespective of the number of encounters with bacteriophages, while the maintenance of CRISPR immunity appears low-cost, but its infection-dependent cost is likely to be much higher. It also seems unlikely that inducible mechanisms would be selected during the relatively short duration of most bacteriophage treatments (days to weeks). In the present study, we observed the in vivo selection of a bacteriophage resistance phenotype in 43.8% (7/16) of patients for which adequate follow up bacterial samples were available for testing. Non-synonymous single-nucleotide polymorphisms (SNPs) or deletions in genes affecting the bacteriophage receptor or coding for a DNA gyrase were assumed to be at the basis of the resistance phenotype in five cases. In two patients, bacterial strains that were no hosts for the applied bacteriophages were selected. In some cases, the in vivo selected bacteriophage-resistant mutants were shown to exhibit re-sensitization to certain antibiotics and reduced virulence in a *G. mellonella* larvae model. The selection of bacteriophage-insensitive bacteria did not prevent the ultimate eradication of the targeted bacterial strains and clinical improvement in four patients.

To date, all BT RCTs evaluated defined bacteriophage products as stand-alone therapies^6^, while bacteriophage-antibiotic synergy is increasingly reported in the literature^65–68^. Based on BT clinical data generated in compassionate use settings, in combination with antibiotic therapy, the ARLG Phage Taskforce recently suggested that BT should be used in conjunction with conventional antibiotics^59^. Correspondingly, we here observed a statistically significant correlation between the eradication of the targeted bacteria and adjunctive standard-of-care antibiotic therapy. In addition, in several of the present 100 cases, it was assumed that the clinical resolution of MDR infections was due to the additive or synergistic effect of various bacteriophage-antibiotic combinations^8,24,26–31,33–35^. In 2016, Chan and colleagues hypothesized, based on in vitro experiments, that the therapeutic use of bacteriophages binding to *P. aeruginosa* efflux pumps could select bacteriophage-resistant isolates with changes in the efflux pump mechanism, causing increased sensitivity to certain chemical antibiotics^69^. In the present study, we demonstrated that the therapeutic use of bacteriophage PT07, predicted to bind to the MexAB-OprM multidrug efflux pump, indeed selected (in vivo) bacteriophage-resistant mutants with changes to the efflux pump mechanism, resulting in increased sensitivity to quinolones. The use of certain bacteriophages in combination with antibiotics seems to re-sensitize bacteria towards antibiotic activity, increasing bacterial killing and decreasing selection of antibiotic-or bacteriophage-resistant clones. To coin a phrase, “Bacteriophages Can Make Antibiotics Great Again”.

Therefore, the PHAGEFORCE study, currently conducted by the present Belgian consortium, was designed as a prospective patient registry aiming to assess the safety and efficacy of personalized BT as an adjunct treatment for difficult-to-treat infections including musculoskeletal infections, chronic rhinosinusitis, sepsis, cystic fibrosis and bronchiectasis^20^. Eligible patients receive either BT alongside standard-of-care antibiotic treatment (BT group) or standard antibiotic treatment alone (Control group), based on the availability of bacteriophages targeting the patient’s infecting bacterial strain(s).

However, caution is warranted, as it is known that certain antibiotics can interfere with bacteriophage lytic activity^70^. While we observed in vitro synergy of colistin and bacteriophage PNM in BT of a *P. aeruginosa* transplant liver infection^27^, colistin and bacteriophage M1 did not show in vitro synergy in BT of *K. pneumoniae* fracture-related infection^8^. We also observed moderate in vitro antagonism of bacteriophage ISP with the antibiotic rifampicin^28^. Rifampicin’s bactericidal properties are based on its inhibition of bacterial RNA polymerases, and its inhibitory effect on some bacteriophages’ RNA synthesis was documented decades ago^71^. Since the concerning bacteriophage (ISP) does not encode for its own RNA polymerase, it relies on the RNA polymerase of its bacterial host for transcription and rifampicin’s mode of action would thus prevent the transcription of ISP’s genome, resulting in an antagonistic effect. It might thus be advised to measure potential synergy, additivism, or antagonism for the proposed combinations of bacteriophages, antibiotics, and the targeted bacterial strain, prior to their clinical application^65^, and this awaiting the availability of more data that might help us predict bacteriophage-antibiotic interactions and thus select the most potent combinations.

In 1922, shortly after he had discovered bacteriophages, Félix d’Hérelle studied the interactions between bacteriophages and immune cells^72^. Since then, a considerable body of experimental data has accumulated showing that bacteriophages can substantially affect immune system cells both in vitro and in vivo, and it has been assumed that anti-bacteriophage antibodies appearing over the course of BT could decrease the lytic activity of bacteriophages and cause therapeutic failure^72^. Consequently, the use of the same bacteriophage(s) for several weeks was discouraged in the former Soviet Union^73^.

In 2014, Łusiak-Szelachowska and colleagues showed that bacteriophage neutralization by sera of patients receiving BT depended on the route of bacteriophage administration and the bacteriophage type, while the induction of anti-bacteriophage activity of sera during BT did not necessarily exclude a favorable BT outcome^74^. In 2017, the same research group suggested that the hampering effect of bacteriophage-recognizing antibodies on the clinical outcome of BT not only depends on the bacteriophage administration route, but also on the dose and frequency, and may not be highly relevant to oral or topical applications, or to low-dose applications, which were shown to elicit weaker or even no immunizations^75^. Given that these studies indicate that antibodies are not formed within a few days, the emergence of bacteriophage-specific antibodies seems less relevant when short treatment courses, in contrast to prolonged courses, or when repeated treatments with the same bacteriophages boost the immune response. The ARLG Phage Taskforce, which advocates using the highest tolerated bacteriophage dose and endorses intravenous bacteriophage administration for infections that involve organs or systems in which bacteriophages have been shown to achieve titers, suggests considering measurement of neutralizing antibodies during prolonged courses of BT^59^. Recently, Dedrick et al. reported on the development of neutralizing antibodies after two months of intravenous BT, which led to treatment failure in an immunocompetent patient with bronchiectasis and *Mycobacterium abscessus* pulmonary infection^76^. In the present study, we also assessed the bacteriophage immune neutralizing effect of patient serum samples against the applied bacteriophages. Since it is assumed that in a serum sample only bacteriophage-neutralizing immunoglobulins could be responsible for a time-dependent loss of bacteriophage titer, this practice indirectly detects bacteriophage-specific antibodies. We observed bacteriophage immune neutralization in five of thirteen evaluated cases, and as could be expected, it emerged 6-35 days after BT initiation, always involved invasive bacteriophage administrations (intravenous and/or intralesional applications), and did not prevent clinical improvement and bacterial eradication in four of five patients. In one patient, a cystic fibrosis patient with an *M. abscessus* lung infection, who was unsuccessfully treated with a bacteriophage (8UZL) administered via nebulization for three weeks and intravenously for ten days, 8UZL serum neutralization emerged seven days after BT initiation, and may thus have hampered BT.

The clinical application of purified bacteriophage preparations is generally considered to be safe, irrespective of the administration route, with a low incidence of adverse events^5^. Correspondingly, only mild to moderate adverse events, possibly related to the bacteriophage product, were reported in 7% (7/100) of the present BT cases.

We acknowledge that our analysis, involving 100 severely ill patients for whom BT was a salvage therapy and our primary aim was to help these patients, has intrinsic limitations. No control groups, blinding, or randomization were put in place and different medical specialties and infection types were involved. Evaluation of safety and efficacy was not based on pre-defined standardized tests, but on the judgment of the treating physicians, and although they were all very experienced, this introduces a certain subjectivity. Then again, it is difficult to evaluate a personalized approach using a classical clinical trial format. Notwithstanding the above-mentioned limitations, we consider that this case series provides key insights that are not only valuable for the treatment of last resort patients, but also for the design of prospective clinical trials such as the PHAGEFORCE study^20^. We confirmed the safety profile of BT and the advantages of combining BT with standard-of-care antibiotic therapy. Samples allowing supportive tests in 21 patients, in view of better treatment management, shed more light on some BT issues such as the in vivo selection of bacteriophage resistance, bacteriophage-antibiotics synergy, and bacteriophage immune neutralization.

Only 27 of the 100 BT cases discussed in this report were published to date, most of them involving complex infections, highlighting some key findings, but most of them leading to a successful outcome. This means that ultimately only about a quarter of the 100 BT cases would have been made public were it not for this publication. The tendency of authors to submit, and of journals to accept, predominantly manuscripts with positive results, was shown to lead to publication bias with positive findings being reported more often, and more quickly, than negative trials^77^. This is most likely also true for the BT field and is an often-cited criticism. Yet, well-documented negative cases could correct misconceptions and prevent senseless repetitions in therapy as well as in clinical trials. The present publication ensures that also our less successful BT cases are reported.

Antimicrobial resistance is predicted to become the leading cause of death worldwide. We demonstrated that it is feasible to set up a flexible and safe development and production pipeline that provides personalized bacteriophage preparations for use in patients with difficult-to-treat bacterial infections. Treating physicians considered BT, as facilitated by the Belgian consortium, to have been beneficial (clinical improvement) for 77 out of 100 patients. In addition to providing immediate assistance to salvage patients, we confirmed and documented the in vivo selection of bacteriophage resistance, even when three bacteriophages targeting three different receptors in one bacterial strain were applied, and a possible emergence of bacteriophage immune neutralization after about one week of invasive BT. The tailored BT approach, as facilitated by the Belgian magistral phage medicine framework, is low-priced and fast compared to the conventional drug licensing pathway and could avoid evolutionary pressures on bacterial populations that might result in an important selection, spread (to clinical settings) and persistence of bacteriophage resistance^68^. As such, personalized BT seems the best way to ensure the sustainability and long-term efficacy of BT, avoiding the past mistakes with antibiotics^68^.

In conclusion, we present an alternative BT paradigm, that is the use of personalized bacteriophages as additive to standard-of-care antibiotics, which overall significantly improved the eradication rate of the targeted bacteria in the considered patient population. The present study can assist healthcare providers, competent authorities, and policymakers with making informed decisions about the use of bacteriophages in difficult-to-treat infections. Finally, it can be useful for designing future controlled clinical trials that are urgently needed to assist the emerging BT field.

## Methods

### Study design and patients

We reviewed the first 100 consecutive BT cases facilitated by a Belgian consortium between January 1, 2008 and April 30, 2022. Within this consortium, the QAMH coordinated most BT cases, selecting and producing bacteriophages, and suggesting BT protocols, while KU Leuven performed supporting genomic analyses of bacteriophages under consideration and of bacterial genomes, and Sciensano controlled the quality and safety of individual bacteriophage preparations.

Physicians requesting BT with QAMH bacteriophage preparations for their patients submitted a BT request to the Phage Therapy Coordination Centre (PTCC) of the QAMH. The PTCC procedure for selecting patients for BT is depicted in Extended Data Figure 1 and is largely determined by clinical need, regulatory approval, and the availability of bacteriophages targeting the infecting bacteria. Clinical applications were performed by, and under the responsibility of, Bacteriophage Therapy Providers* in several hospitals in Belgium and abroad. Demographic and clinical data were collected through the patients’ treating physicians. Written informed consent for BT was obtained from the involved patients or their legal representatives according to local provisions. Where warranted, local ethical committee approval for BT was obtained. According to EU Regulation No 536/2014 (Clinical Trials Regulation)^78^, its transposition to Belgian Law, and per advice of the Leading Ethical Committee of the “Universitair Ziekenhuis Antwerpen” and the “Universiteit Antwerpen” (ID 3644), the present retrospective non-interventional analysis of an existing and de-identified BT database was not considered as an experiment on the human person and did not require a dedicated informed consent. The observational study protocol was registered on ClinicalTrials.gov (Study BT100, ID: NCT05498363).

### Manufacture of bacteriophage active pharmaceutical ingredients (APIs)

Bacteriophages were isolated and characterized by the QAMH or were sourced from Bacteriophage Donors**. Bacteriophage suspensions were produced in accordance with the guidelines provided by the bacteriophage API monograph^19^, and the methods described by Merabishvili and colleagues^79^, with some modifications. Bacteriophage stocks were prepared using the double agar overlay method with minor modifications. Three to six milliliters of bacteriophage lysate containing 10^3^-10^5^ PFU of bacteriophages were added to a sterile 15 mL Falcon tube (Greiner Bio-One, Vilvoorde, Belgium) and complemented with 0.2 mL of a bacteriophage-sensitive bacterial suspension (end concentration of 10^8^ CFU/mL) and lukewarm medium [Select Alternative Protein Source (APS) Lysogeny Broth (LB), Tryptic Soy Broth (TSB), or TSB + 0.5% glycerol (all purchased from Becton Dickinson, Erembodegem, Belgium)] with 0.6% top agar (VWR International, Leuven, Belgium), to a total volume of 12 mL. This mixture was plated onto a square (12 × 12 cm) Petri dish (Greiner Bio-One) filled with a bottom layer of APS LB, TSB, or TSB medium + 0.5% glycerol (all Becton Dickinson) and 1.5% agar (VWR International), and incubated at 32 °C (for *E. coli*, *K. pneumoniae*, and *P. aeruginosa*) or 37 °C (for all the other bacterial species) for 16 h or 48 h (for *M. abscessus*). The top agar layer was scraped off using a sterile L-shaped rod (Sigma Aldrich, Overijse, Belgium), transferred to a sterile 50 mL sterile Falcon tube (Greiner Bio-One), and centrifuged for 20 min at 6,000 *g* using a Sorvall™ Legend™ centrifuge (Thermo Fisher Scientific, Dilbeek, Belgium). The supernatant was aspirated using a sterile 30 mL syringe (BD Plastipak, Becton Dickinson) with an 18 G sterile needle (BD microlance 3, Becton Dickinson) and filtered sequentially by a 0.45 µm and a 0.22 µm polyethersulfone (PES) Millex^®^-Gp membrane syringe filter (Merck, Darmstadt, Germany) or using a vacuum filter system (Nalgene, Thermo Fisher Scientific). The bacteriophage suspension was centrifuged for 90 min at 35,000 *g* (40,000 *g* for podoviruses) using a Sorvall™ Legend™ centrifuge (Thermo Fisher Scientific). The resulting bacteriophage pellet was diluted in ten times less Dulbecco’s phosphate buffered saline without Calcium and Magnesium (DPBS, Lonza, Verviers, Belgium) than the initial bacteriophage suspension, and the pellet was left to dissolve overnight at 4 °C. The bacteriophage suspension was further diluted to a final concentration of 10^9^– 10^10^ PFU/mL using DPBS (Lonza). The diluted bacteriophage suspension was filtered using a 0.22 µm PES Millex^®^-Gp membrane syringe filter (Merck) and subsequently purified from endotoxins using the commercially available kits EndoTrap^®^ Blue (Lonza, Verviers, Belgium) or EndoTrap^®^ HD (Lionex, Braunschweig, Germany), according to the instructions of the manufacturer. One column was utilized per 50 mL of bacteriophage suspension. Endotoxin purified bacteriophage suspensions were filtered using medical grade 0.22 µm Polyvinylidene difluoride (PVDF) Millex^®^-Gp syringe filters (Merck) and collected into sterile 125 or 500 mL PETG Nalgene^®^ bottles (Thermo Fisher Scientific). The final titer of each thus obtained bacteriophage API was 10^9^–10^10^ PFU/mL.

### Quality and safety of bacteriophage APIs

Sciensano controlled the quality and safety of the bacteriophages. In accordance with the bacteriophage API monograph^19^, this control was implemented on two levels (https://www.sciensano.be/en/control-and-safety-assessment/safety-therapeutic-bacteriophage-preparations).

First, a genetic control was performed to check the safety of the bacteriophage to be used in human therapy. For this purpose, genomic DNA of the bacteriophages and their bacterial hosts were isolated and purified using respectively a MagCore^®^ Viral Nucleic Acid and a MgC Bacterial DNA Kit™ with a 60 μL elution volume (Atrida, Amersfoort, The Netherlands), following the manufacturer’s instructions. Sequencing libraries were constructed using the Illumina Nextera XT DNA sample preparation kit and sequenced on an Illumina MiSeq instrument with a 250 bp paired-end protocol (MiSeq v3 chemistry, Illumina, San Diego, CA, USA). Trimming of short reads was performed with Trimmomatic (version 0.32)^80^. Additionally, for bacterial production strains, long-read sequencing was performed using ONT’s Rapid barcoding kit SQK-RBK004 and a MinION flow cell (v9.4.1), according to the manufacturer’s instructions. Super High Accuracy base calling was performed using Guppy (v6.0.1) (Oxford Nanopore Technologies–ONT, Oxford, UK), and hybrid assemblies were generated using Unicycler (v0.4.7)^81^. For bacteriophages, genome assembly was performed using SPAdes (Galaxy Version 3.15.4+)^82^, after which the genome was annotated using Prokka (v1.14.6)^83^ with assistance of the PHROGS database (https://phrogs.lmge.uca.fr/). To detect undesired genes associated with antibiotic resistance or virulence, the complete bacteriophage genome was submitted to the NCBI (National Center for Biotechnology) blastn web interface (https://blast.ncbi.nlm.nih.gov/Blast.cgi) for a similarity search in different databases (ARG-ANNOT, CARD, ResFinder, and VFDB). Prophage induction was searched by mapping sequencing reads of the production batch to the bacterial production host genome and looking for significantly increased coverage in predicted prophage positions (determined using PHASTER^84^ and Prophage Hunter^85^).

Second, Sciensano analyzed various parameters of each production lot of each bacteriophage API. Bacteriophage identity was determined using DNA extraction and genome sequencing as described above. The potency of the lot was verified using classical double agar dilutions, in triplicate. The bioburden and pH of each lot was assessed as described in European Pharmacopoeia (Ph. Eur.) chapters 2.6.12 and 2.2.4, respectively. Endotoxin levels were measured using the Limulus Amebocyte Lysate (LAL) Test, according to Ph. Eur. chapter 2.6.14. A certificate allowing the bacteriophage API to be used in pharmaceutical (magistral) preparations was provided by Sciensano upon successful completion of this two-tiered procedure.

### Selection of adequate bacteriophages for therapy

The patients’ infecting bacteria were sent to the PTCC, and their bacteriophage susceptibility was determined. Susceptibility of bacterial strains towards the available bacteriophage cocktails or APIs was tested using the spot test as described by Elizabeth Kutter^86^. Fresh overnight cultures of the patient’s bacterial strains were added to lukewarm (46 °C) media containing 0.6% agar (top agar) and poured onto square (12 × 12 cm) Petri dishes (Greiner Bio-One) containing media with 1.5% agar (bottom agar). Different culture media were used, according to the considered bacterial species. Media were purchased from Becton Dickinson and agar from VWR International. Droplets (10 µL) of serial dilutions of each of the considered bacteriophage solutions were spotted on the top agar layer. Petri dishes were incubated overnight at 32 or 37 °C, according to the considered bacterial species. The next day, the lysis zones produced by active bacteriophages in the bacterial lawn were examined and classified as confluent lysis (4+), semi-confluent lysis (3+), opaque lysis (2+), separate plaques (+), or no activity (-). Next, for bacteriophages producing clear lysis zones, EOP was defined, as described by Kutter^86^. The EOP for the patient’s bacterial strain was calculated by comparison with a highly susceptible reference host and was defined as the observed number of PFUs on the patient’s bacterial strain (as determined by the above-described spot test) divided by the observed number of PFUs on the reference bacterial strain. The EOP value obtained with the highly susceptible production host strain was considered as EOP = 1.0. In case the picture was unclear (e.g. opaque lysis zones) and the results difficult or un-interpretable, the double agar overlay method was used to determine the PFUs on the patient’s strains and the bacteriophage production host, as described above, to define EOP more precisely. When the activity of the bacteriophages was still difficult to assess using the above-mentioned methods based on solid media, liquid broth cultures were used to assess bacteriophage activity, using the OmniLog^®^ system (Biolog, Hayward, CA, USA). Bacterial respiration was measured without and with bacteriophages. Experiments were performed in 96-well plates (Thermo Fisher Scientific) in a final volume of 200 µL of LB or TSB medium (Becton Dickinson), supplemented with 100-fold diluted tetrazolium dye mix A or H (Biolog). Bacterial cells were inoculated at a concentration of 10^5^ CFU/well, calculated based on optical density (OD) at 600 nm and validated using a classical plate culture method. Bacteriophages were added at an MOI range of 100–0.0001, as calculated on the propagation host. Plates were incubated at a bacterial species-specific temperature (32 or 37 °C) for 72 **h**, and the color change caused by reduction of the tetrazolium dye due to bacterial respiration (during growth) was recorded every 15 min by the OmnilLog^®^ system. The results were analyzed with Biolog Data Analysis software (v1.7) and data was exported into Microsoft Excel files.

For the treatment of patients, bacteriophages with the highest EOPs were selected. Since April 2022, when more than one bacteriophage showed adequate in vitro activity, the overall activity of the bacteriophage combinations was analyzed using the OmniLog^®^ system, as described above. When synergistic or additive effects were observed, the concerned bacteriophage combinations were recommended for clinical use.

### Pre-adaptation of bacteriophages

When the observed bacteriophage susceptibility was deemed too low for therapeutic application, and if time and resources permitted, bacteriophages were pre-adapted to increase pathogen clearance and to reduce bacteriophage resistance evolution^54–56^. According to the guidelines of the Ministry of Health of the USSR and the empirical experience of the Eliava Institute, adequate bacteriophage cocktails should cause stable lysis – that is without the emergence of bacteriophage-insensitive bacterial mutants – of the target bacteria, in liquid medium, for a prolonged time (typically 48 h), and at a multiplicity of infection MOI of 0.0001–0.00001 and bacterial concentrations of 10^6^ CFU/mL^87–90^. For individual bacteriophages, MOIs ≤ 1.0 are deemed appropriate. To obtain these bacteriophage virulence and bacterial regrowth suppression thresholds, the (modified) Appelmans method was applied for the pre-adaptation of bacteriophages on bacterial strains, as previously described^91^. To a 15 mL Falcon tube (Greiner Bio-One) were added: 4.5 mL of LB or TSB medium (Becton Dickinson), 0.5 mL of tenfold dilutions of the considered bacteriophage, and a volume of either the patient’s bacterial strain or a pre-production panel of collected “problematic” bacterial strains, to obtain a final concentration of 10^6^ CFU/mL. The tubes were incubated at a bacterial species-specific temperature (32 or 37 °C) for 48 h. Bacterial growth and bacteriophage activity were monitored by OD measurement at 600 nm, using a Lambda 12 UV/VIS spectrometer (Perkin Elmer, Waltham, MA, United States) after 24 and 48 h of incubation and compared to two negative controls (bacteriophage only and LB or TSB medium only) and a positive control (bacteria only). The tube with the highest bacteriophage dilution showing an OD_600_ value similar to the negative controls was selected and chloroform was added to a final concentration of 2.0% (vol/vol). The tube was shaken and incubated for at least 2 h at 2–8 °C. After incubation, the upper phase (without chloroform) was aspirated using a sterile 30 mL syringe (BD Plastipak, Becton Dickinson) with an 18 G sterile needle (BD microlance 3, Becton Dickinson) and filtered using a 0.45 µm or a 0.22 µm PES Millex^®^-Gp membrane syringe filter (Merck). The obtained bacteriophage lysate underwent several (at least three) of the above-described passages until adequate virulence and resistance suppression levels were obtained.

### Bacteriophage preparation stability

The stability of the bacteriophage APIs was monitored by determining their titer at 2–8 °C on a monthly basis. Bacteriophage APIs, with titers of 10^9^–10^10^ PFU/mL, retained their activity for at least one year^92^. One or more bacteriophage APIs can be diluted and/or mixed with a carrier (e.g. an isotonic intravenous solution or a hydrogel) into a magistral preparation, under the supervision of a hospital pharmacist, and according to the provisions of a medical prescription provided by the patient’s treating physician. Diluting and mixing various bacteriophages are events that can compromise their stability^92,93^, and experiments showed that, in general, magistral preparations are best used within one week after their manufacture.

### Bacteriophage therapy protocols

The PTCC suggested BT protocols based on the application instructions of the Ministry of Health of the USSR^21–23^ and the experiences of the Eliava Institute (personal communications), some of which can be found in the leaflets of their BT products.

For nebulization of bacteriophage preparations, vibrating mesh type nebulizers were advised because they were shown to induce less titer loss, due morphological damage, than air-jet nebulizers^94,95^, and for bone and orthopedic prosthesis infections we advised to use a pigtail catheter or another draining device for rinsing the wound cavities prior to bacteriophage application and for the actual administration of bacteriophages^29^. For topical application, we advised to mix the bacteriophages with an adequate hydrogel^93^.

### Supporting assays

In addition to bacteriophage susceptibility testing, three BT supportive tests were offered, without obligation, to the Bacteriophage Therapy Providers**, to allow for improved BT management.

#### In vivo selection of bacteriophage resistance

The in vivo selection of bacteriophage resistance was monitored using sequential bacterial samples isolated during BT. Bacteriophage susceptibility was evaluated using the methods described earlier. When decreased bacteriophage sensitivity was observed, the isolate’s genome was sequenced and analyzed to determine the clonality of the isolate (compared with the pre-BT isolate) and to investigate the genetic background for the observed bacteriophage resistance phenotype. For genome sequencing, the method described in Eskenazi *et al*. (2022)^8^ was followed with some deviations: for nanopore processing, Guppy (v6.3.8) (Oxford Nanopore Technologies) (basecalling, demultiplexing) and Porechop (v0.2.4) (barcode clipping) (https://github.com/rrwick/Porechop) were used. Genomes were assembled with Unicycler (v0.4.8)^81^ and SNP variants were called using Snippy (v4.6.0) (https://github.com/tseemann/snippy). For genome annotation and visualization, EggNOG-mapper (v2.1.8)^96^, mobileOG-db (v1.1.2)^97^, Phigaro (v.2.3.0)^98^, circos (v.0.69.8)^99^, and GC-profile^100^ were used. A pan-genome analysis using Roary (v3.13.0)^101^ from annotated genomes (Prokka v1.14.6)^83^ was performed to create a maximum likelihood phylogenetic tree using core alignment in fasttree (v2.1.10) visualized with iTOL^102^. For Multi-locus sequence typing (MLST), genomes were scanned against PubMLST schemes, including ST111 (O12-1709), ST357 (B14130), ST235 (NCGM2), ST1233 (PcyII-10) PAO1 (ST549) and ATCC 27853 (ST155) as representative genomes/STs.

#### In vitro bacteriophage-antibiotic interactions

In vitro bacteriophage-antibiotic interactions were analyzed using the initial bacterial isolates obtained prior to the start of BT. Bacterial respiration was measured using the OmniLog^®^ system (Biolog). The growth kinetics of the targeted bacterial pathogens were assessed in the presence of the bacteriophages only, the relevant antibiotics (to be used concomitantly) only, and bacteriophage-antibiotic combinations. Experiments were performed in triplicate (biological replicates) in 96-well plates (Thermo Fisher Scientific) in a final volume of 200 µL of LB or TSB medium (Becton Dickinson) supplemented with 100-fold diluted tetrazolium dye mix A or H (Biolog). Bacterial cells were inoculated at a concentration of 10^5^ CFU/well, calculated based on OD_600_ measurements and validated using a classical plate culture method. Antibiotics and bacteriophages were added at sub-MIC concentrations and MOIs ≤ 1.0 (calculated on the propagation host), respectively. The titers of the bacteriophages were confirmed after each experiment using the classical double agar overlay method. Plates were incubated at 37 °C for 72 h and the color change caused by reduction of the tetrazolium dye due to bacterial respiration (during growth) was recorded every 15 min by the OmnilLog^®^ system. The results were analyzed with Biolog Data Analysis software (v1.7) and data was exported into Microsoft excel files.

#### Bacteriophage immune neutralization

The possible emergence of bacteriophage immune neutralization, or the ability of the patient’s serum to neutralize therapeutic bacteriophages, was evaluated according to Adams^103^, with some modifications. Whole blood samples were collected prior to BT initiation and at various time points during and after bacteriophage application. Blood was allowed to clot for at least 30 min in a vertical position and then centrifuged in a swinging bucket rotor for 10 min at 2,000 *g* at room temperature. The obtained serum samples were stored at −80 °C ± 5 °C. To assess the effect of the serum samples on bacteriophage lytic activity, 0.9 mL of 1:100 diluted sera were mixed with 0.1 mL of the bacteriophage suspension at a concentration of 2 x 10^7^ PFU/mL and incubated for 30 min at 37 °C. Bacteriophage lytic activity (titer) was determined before and after incubation with the patient’s serum samples using the double agar overlay plaque assay (as previously described). Comparison of pre- and post-incubation lytic activity allowed for the determination of the proportion of neutralized bacteriophages. Each serum sample was tested in triplicate.

#### Galleria mellonella virulence assay

Ten *P. aeruginosa* isolates [Pa30 (Is1), Pa30 (Is3), Pa54 (Is1), Pa54 (Is4), and Pa91 (IS1-6)] were grown in LB broth (Becton Dickinson) to an OD_600_ of 0.25–0.35. One milliliter of the bacterial cultures was centrifuged and re-suspended in sterile DPBS (Lonza). *G. mellonella* larvae were grouped in batches of 10 (standardized for weight) and then injected in the hindmost proleg with a 10 µl aliquot of 10^-5^ dilutions (± 10 CFUs) of the washed bacterial cultures. After infection, the larvae were incubated in the dark at 37 °C. Health status was monitored every 24 h and compared to DPBS (Lonza) injected controls.

### Clinical outcome

Clinical improvement, eradication of the targeted bacterium, and the advent, severity and duration of adverse events or reactions were assessed by the treating physician.

### Data collection

Prior to BT, demographic and clinical data were collected through a medical form, which was completed by the Bacteriophage Therapy Providers*. The medical doctor’s BT prescription, information regarding the applied bacteriophage product and its administration route, dosage, duration, and information with regard to possible concomitant (antibiotic) treatments were also recorded. The “phagograms”, reporting on the evaluation of the bacteriophage susceptibility of the patient’s bacterial isolates sampled before and sometimes during treatment were also archived. If the bacteriophage treatment was performed in a hospital, a clinical follow-up form, requesting information about the clinical outcome (incl. possible adverse events and reactions), was completed by the treating physician and the nursing team and sent to the PTCC. In case of ambulatory BT, clinical follow-up information was collected directly from the patients. All demographic, bacteriophage product, and clinical data were recorded in a REDCap (Research Electronic Data Capture) designed database^104^.

### Definitions

In accordance with the guidelines of an international expert proposal for interim standard definitions for acquired resistance, multidrug resistance (MDR) was defined as acquired non-susceptibility to at least one agent in three or more antimicrobial categories, extensive drug resistance (XDR) as non-susceptibility to at least one agent in all but two or fewer antimicrobial categories, and pandrug resistance (PDR) as non-susceptibility to all agents in all antimicrobial categories^105^. The term “usual drug resistance” (UDR) was used to describe isolates that are not fully susceptible, but could nonetheless be readily treated (at least based on the *in vitro* susceptibility assays) using standard therapies^106^. If an infection persisted for more than six months, it was considered a “chronic infection”.

Clinical improvement was defined as the improvement of at least one symptom associated with the bacterial infection, as assessed by the treating physician. Eradication of the targeted bacterium was defined as the absence of the originally targeted causative agent of the bacterial infection in culture, or when the patient’s treating physician concluded, based on a follow up survey, that the patient was freed of the targeted bacterial pathogen. The period between the start of BT and the evaluation of the clinical outcome varied according to the treating physician and the indication, and ranged from one month to one year, the latter for difficult-to-treat bone infections.

### Statistical methods

The following variables were analyzed for 92 of the 100 patients (for which a complete data set was available): eradication of the targeted bacteria, clinical improvement, concomitant use of antibiotics, antibiotic resistance profile of the target bacteria, adverse reactions, and the clinical setting (ambulatory treatment or hospitalized). All these variables were binary categorical. In addition, the 14 infection types and 21 bacterial species targeted by BT were monitored on nominal categorical scales. Age and gender were analyzed on numeric scales. The statistical analysis was conducted using the statistical software environment SAS (v9.4) (Cary, NC, USA). We used a stepwise selection procedure on a reduced dataset (Supplementary Table 2) to determine the most informative variable in the dataset, with the variable “Eradication (ERADIC)” as response variable of our logistic regression model. The probability modeled is ERADIC = “Yes” (i.e. successful eradication). The data presented in Fig. 1 (patient population characteristics) and Fig. 4 (bacteriophage immune neutralization) were analyzed using R (v4.3.0) (https://www.R-project.org/) ^107^ and visualized with the following packages: tidyverse (v2.0.0)^108^, UpSetR (v1.4.0)^109^, ggmap (v3.0.2)^110^, and rnaturalearth (R package version 0.3.2.9000)^111^. The log-rank test with Bonferroni correction for multiple comparisons (GraphPad V 0.5.1) was used for *G. mellonella* survival curve comparisons.

## Data availability

Detailed clinical protocols, results, and additional data are available in the manuscript and in Supplementary Tables 1 and 2. The protocol for the retrospective, observational study is available at: https://clinicaltrials.gov/ct2/show/NCT05498363?term=NCT05498363&draw=2&rank=1. The bacteriophage genome sequences can be retrieved in the GenBank database under the accession codes listed in Extended Data Table 2. The genome data of the bacterial isolates can be accessed via the NCBI BioProject PRJNA975428. The authors declare that all other data supporting the findings of this study are available within the article.

## Acknowledgements

CC, GS, MM, and TG were supported by the Royal Higher Institute for Defence, Brussels, Belgium. This work would not have been possible without the support of Elizabeth (Betty) Kutter, Mario Vaneechoutte, Revaz Adamia, Alain Vanderkelen, Pierre Neirinckx, and Geert Laire.

## Author contributions

MM, GS, JG, NC, MK, CC, TG, and Bacteriophage Donors* isolated, characterized and made bacteriophages available for clinical use. MM, GS, and JG performed the *in vitro* matching (phagogram) of bacteriophages to the patients’ infecting bacteria. MM and BVN performed the bacteriophage-antibiotic interaction assays. MM, GS, and JG produced the bacteriophage APIs and a substantial part of the magistral bacteriophage preparations. MDJ, P-JC, JW, CL, SG, RL, and Bacteriophage Donors* sequenced and analyzed the genomes of the selected bacteriophages and bacteria. SG designed and performed the *Galleria mellonella* experiments. MM, BVN, and JO performed the bacteriophage immune neutralization assays. MDJ, P-JC, J-PD, and GV designed the quality management system, and performed and validated the quality control of the bacteriophage preparations. SD, JO, BVN, NC, MK, PS, and J-PP screened the BT requests, selected and coordinated BT cases, including the suggestion of BT protocols. SD, JO, BVN, NC, MK, J-PP, and Bacteriophage Therapy Providers** facilitated or performed BT cases. SD, AS, EV and J-PP collected and summarized all relevant clinical and epidemiological data. ES, CL and J-PP performed the statistical analysis. CL, SG, and J-PP drafted the figures and tables. J-PP prepared the first draft of the manuscript. All authors revised the final version and agreed to the submission of the final version.

## Correspondence and requests for materials

should be addressed to Jean-Paul Pirnay.

## ^*^Bacteriophage Therapy Providers

Kim Win Pang^2^, Willem-Jan Metsemakers^7^, Dimitri Van der Linden^9^, Olga Chatzis^9^, Anaïs Eskenazi^10^, Angel Lopez^11^, Adrien De Voeght^12^, Anne Françoise Rousseau^13^, Anne Tilmanne^14^, Daphne Vens^15^, Jean Gérain^16^, Brice Layeux^17^, Erika Vlieghe^18^, Ingrid Baar^19^, Sabrina Van Ierssel^19^, Johan Van Laethem^20^, Julien Guiot^21^, Sophie De Roock^22^, Serge Jennes^22^, Saartje Uyttebroek^23^, Laura Van Gerven^23^, Peter W. Hellings^23^, Lieven Dupont^24^, Yves Debaveye^25^, David Devolder^26^, Isabel Spriet^27^, Paul De Munter^28^, Melissa Depypere^29^, Michiel Vanfleteren^30^, Olivier Cornu^31^, Stijn Verhulst^32^, Tine ^32^, Stoffel Lamote^33^, Thibaut Van Zele^34^, Grégoire Wieërs^35^, Cécile Courtin^36^, David Lebeaux^37^, Jacques Sartre^38^, Tristan Ferry^39^, Frédéric Laurent^40^, Kevin Paul^41^, Mariagrazia Di Luca^42^, Stefan Gottschlich^43^, Tamta Tkhilaishvili^44^, Novella Cesta^45^, Karlis Racenis^46^, Telma Barbosa^47^, Luis Eduardo López-Cortés^48^, Maria Tomás^49^, Martin Hübner^50^, Truong-Thanh Pham^51^, Paul Nagtegaal^52^, Jaap Ten Oever^53^, Ghariani Iheb^54^, Joshua Jones^55^, Lesley Hall^56^ and Matthew Young^57^

9 Pediatric Infectious Diseases, Pediatric Department, Cliniques Universitaires Saint-Luc, Université Catholique de Louvain-UCLouvain, Brussels, Belgium.

10 Clinic of Infectious Diseases, Erasme Hospital, Brussels, Belgium.

11 HNO-Zentrum Simmering, Vienna, Austria.

12 Department of Medicine, Division of Hematology, Centre Hospitalier Universitaire de Liège, University of Liège, Liège, Belgium.

13 Department of Intensive Care and Burn Center, University Hospital, University of Liège, Liège, Belgium.

14 Senior Lecturer in Infectious Diseases, Faculté de Médecine, Université Libre de Bruxelles, Brussels, Belgium.

15 Division of Pediatric Infectious Diseases and Infection Prevention and Control, Hôpital Universitaire des Enfants Reine Fabiola, Université Libre de Bruxelles (ULB), Brussels, Belgium.

16 Department of Internal Medicine, Delta Hospital (CHIREC), Brussels, Belgium.

17 Delta Hospital (CHIREC), Brussels, Belgium.

18 18 Department of General Internal Medicine, Infectious Diseases and Tropical Medicine, University Hospital Antwerp, Edegem, Belgium.

19 Department of Critical Care Medicine, University Hospital Antwerp, University of Antwerp, Edegem, Belgium.

20 Department of Internal Medicine and Infectious Diseases, Vrije Universiteit Brussel, Universitair Ziekenhuis Brussel, Brussels, Belgium.

21 Pneumology Department, CHU Liège, Domaine universitaire du Sart-Tilman, Liège, Belgium.

22 Queen Astrid Military Hospital, Brussels, Belgium.

23 Department of Otorhinolaryngology, Head and Neck surgery, University Hospitals Leuven; Experimental Otorhinolaryngology, Rhinology Research, Department of Neurosciences, KU Leuven, Leuven, Belgium.

24 Department of Pneumology, University Hospital Leuven; Respiratory Diseases and Thoracic Surgery, Department of Chronic Diseases and Metabolism, KU Leuven, Leuven, Belgium.

25 Department of Intensive Care Medicine, University Hospitals Leuven; Department of Cellular and Molecular Medicine, KU Leuven, Leuven, Belgium.

26 Pharmacy Department, University Hospitals Leuven, Leuven, Belgium.

27 Pharmacy Department, University Hospitals Leuven; Clinical Pharmacology and Pharmacotherapy, Department of Pharmaceutical and Pharmacological Sciences, KU Leuven, Leuven, Belgium.

28 Department of General Internal Medicine, University Hospitals Leuven; Department of Microbiology, Immunology and Transplantation, KU Leuven, Leuven, Belgium.

29 Department of Laboratory Medicine, University Hospitals Leuven; Laboratory of Clinical Bacteriology and Mycology, KU Leuven, Leuven, Belgium.

30 St-Jozefskliniek Izegem, Izegem, Belgium.

31 Department of Orthopaedic Surgery, University Hospital Saint Luc, Brussels, Belgium.

32 Department of Pediatrics, Antwerp University Hospital, Edegem, Belgium.

33 Department of Intensive Care Medicine, AZ Groeninge, Kortrijk, Belgium.

34 Department of Otorhinolaryngology, UZ Ghent University Hospital, Gent, Belgium.

35 Service de Médecine Interne Générale, Clinique Saint Pierre, Ottignies, Belgium.

36 Service de Dermatologie, Cliniques Saint-Pierre, Ottignies, Belgium.

37 Service de Microbiologie, Unité Mobile d’Infectiologie, AP-HP, Hôpital Européen Georges Pompidou, Paris, France.

38 Laboratoire de biologie médicale, Centre Hospitalier de Valence, Valence, France.

39 Centre de Référence des Infections Ostéo-Articulaires Complexes de Lyon (CRIOAc Lyon), Hospices Civils de Lyon, Lyon, France.

40 Institut des Agents Infectieux - Hospices Civils de Lyon, Centre International de Recherche en Infectiologie, Lyon, France.

41 University Children’s Hospital, University Medical Center Hamburg-Eppendorf, Hamburg, Germany.

42 Department of Biology, University of Pisa, Pisa, Italy.

43 Praxis Cordes & Gottschlich & Heß & Scherl, Rendsburg, Germany.

44 Department of Cardiothoracic and Vascular Surgery, Charité German Heart Center, Berlin, Germany.

45 PhD Course in Microbiology, Immunology, Infectious Diseases, and Transplants (MIMIT), University of Rome Tor Vergata, Rome, Italy.

46 Department of Biology and Microbiology, Riga Stradins University, Riga, Latvia.

47 Department of Pediatrics, Maternal Child Center of the North (CMIN), University Hospital Center of Porto (CHUP), Porto, Portugal.

48 Enfermedades Infecciosas y Microbiología Clínica, Hospital Universitario Virgen Macarena, Sevilla, Spain.

49 Translational and Multidisciplinary Microbiology Group (MicroTM) - Institute of Biomedical Research A Coruña (INIBIC) and Microbiology Deparment of Hospital of A Coruña (CHUAC), University of A Coruña (UDC), A Coruña, Spain.

50 Department of Visceral Surgery, Lausanne University Hospital CHUV, University of Lausanne (UNIL), Lausanne, Switzerland.

51 Service des maladies infectieuses, Département de médecine, Hôpitaux universitaires de Genève, Genève, Switzerland.

52 Department of Otorhinolaryngology and Head and Neck Surgery, Erasmus MC, Rotterdam, The Netherlands.

53 Department of Internal Medicine and Radboud Centre for Infectious Diseases, Radboud University Medical Center, Nijmegen, The Netherlands.

54 Clinique Saint Augustin, Tunis, Tunisia.

55 Edinburgh Medical School: Biomedical Sciences, University of Edinburgh, Edinburgh, UK.

56 Diabetes and Endocrinology, Queen Elizabeth University Hospital, Glasgow, United Kingdom.

57 Diabetes Foot Clinic, Royal Infirmary, Edinburgh, UK.

## ^**^Bacteriophage Donors

Nana Balarjishvili^5^, Marina Tediashvili^5^, Yigang Tong^58^, Christine Rohde^59^, Johannes Wittmann^59^, Ronen Hazan^60^, Ran Nir-Paz^60^, Joana Azeredo^61^, Victor Krylov^62^, Gregory Resch^63^, Shawna McCallin^64^, Matthew Dunne^65^ and Samuel Kilcher^65^

58 College of Life Science and Technology, Beijing University of Chemical Technology, Beijing, China.

59 Leibniz Institute DSMZ, German Collection of Microorganisms and Cell Cultures GmbH, Braunschweig, Germany. 66

60 Israeli Phage Therapy Center (IPTC) of Hadassah Medical Center and the Hebrew University, Jerusalem, Israel.

61 Department of Biological Engineering, University of Minho, Braga, Portugal.

62 Mechnikov Research Institute of Vaccines & Sera, Moscow, Russia.

63 Laboratory of Bacteriophages, Lausanne University Hospital, Lausanne, Switzerland.

64 Department of Neuro-Urology, Balgrist University Hospital, University of Zürich, Zürich, Switzerland.

65 Institute of Food, Nutrition and Health, ETH Zurich, Zurich, Switzerland.

---

**Extended Data Table 1.**
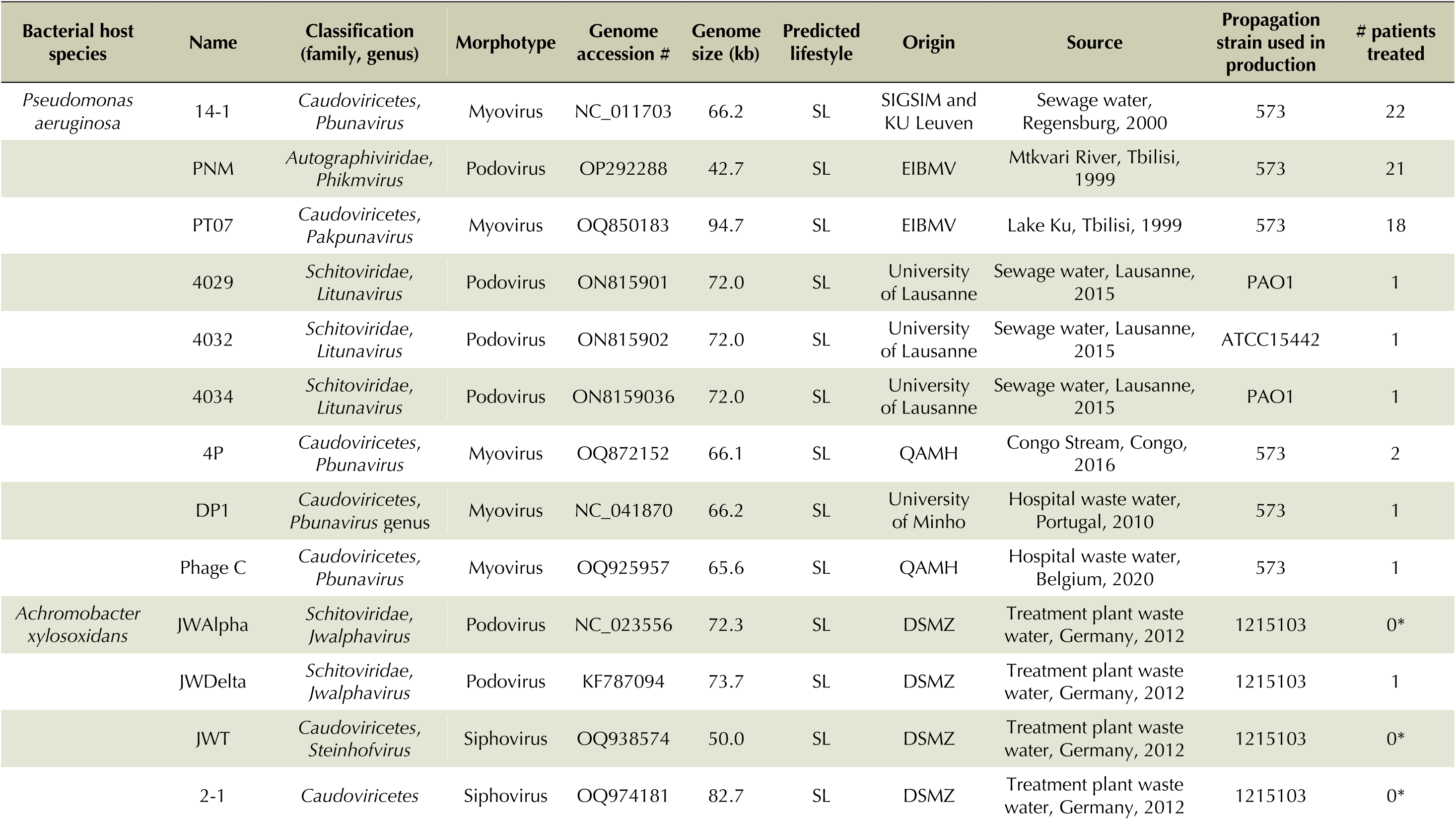
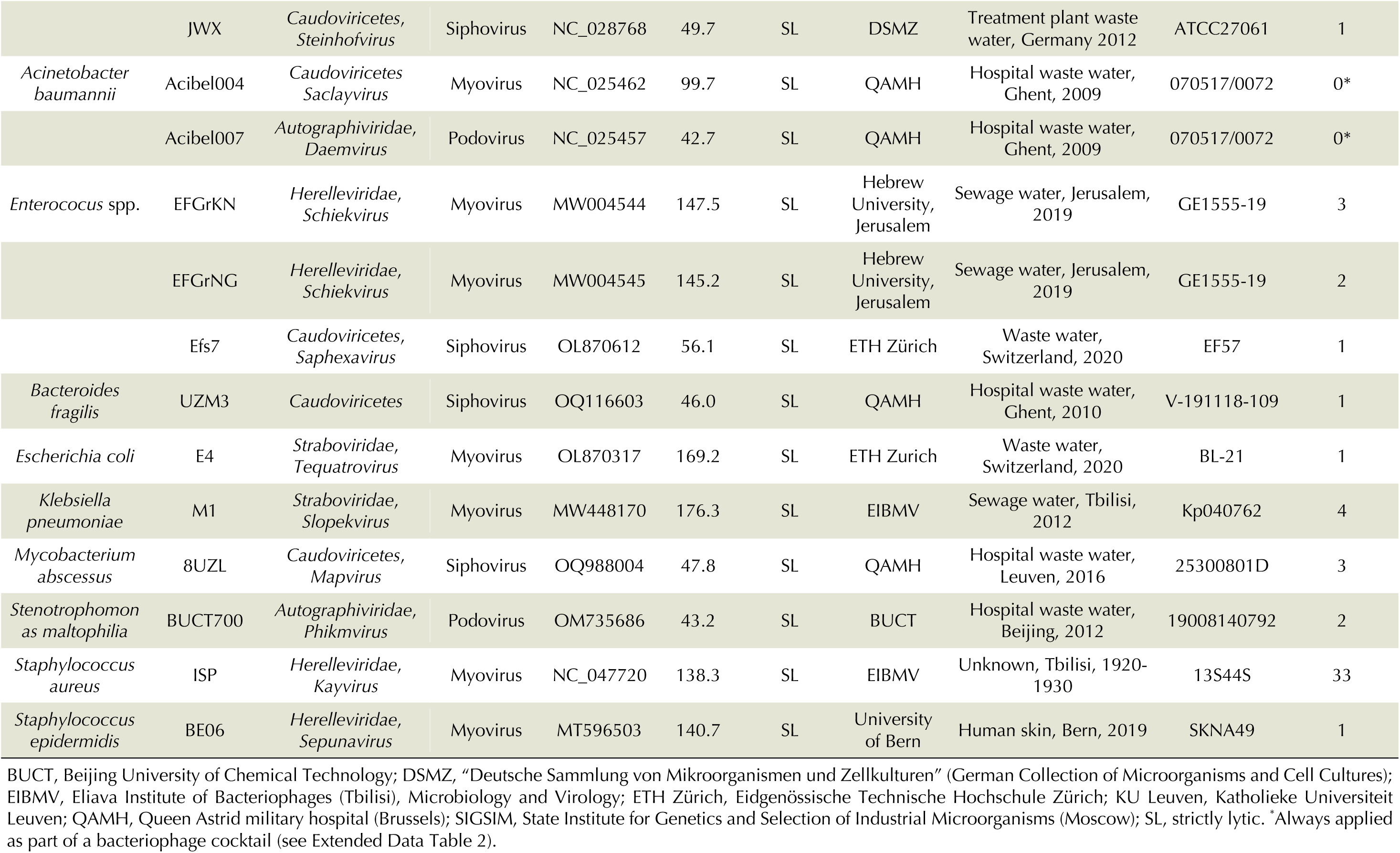
Characteristics of the 26 bacteriophages used, individually or in combination, in the present 100 consecutive bacteriophage therapy cases.

**Extended Data Table 2.**
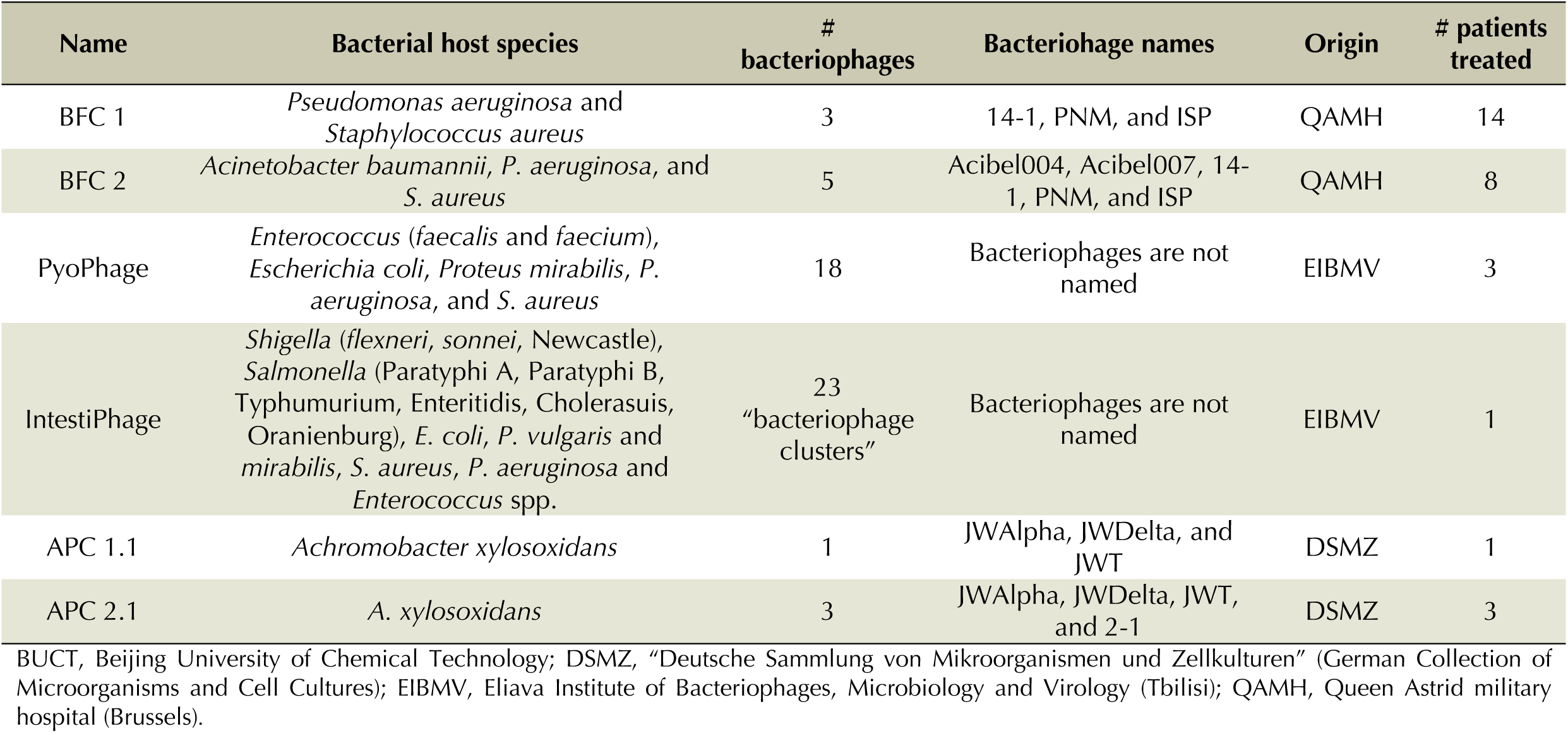
Characteristics of the six defined bacteriophage cocktails used in the present 100 consecutive bacteriophage therapy cases.

**Extended Data Table 3.**
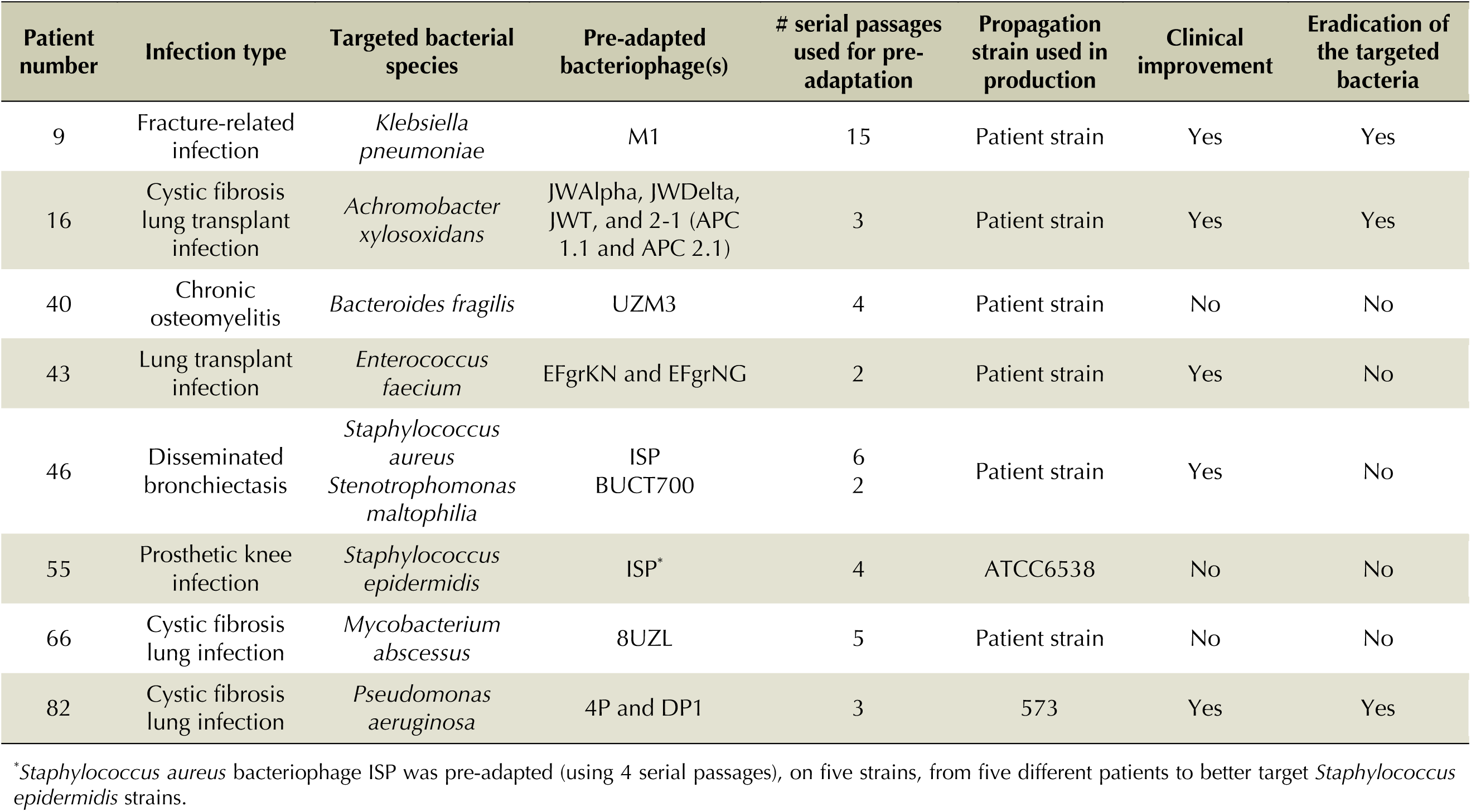
Characteristics of the bacteriophage therapy cases that necessitated pre-adaptation of bacteriophages.

| Patient number | Infection type | Targeted bacterial species | Pre-adapted bacteriophage(s) | # serial passages used for pre-adaptation | Propagation strain used in production | Clinical improvement | Eradication of the targeted bacteria |
| --- | --- | --- | --- | --- | --- | --- | --- |
| 9 | Fracture-related infection | *Klebsiella pneumoniae* | M1 | 15 | Patient strain | Yes | Yes |
| 16 | Cystic fibrosis lung transplant infection | *Achromobacter xylosoxidans* | JWAlpha, JWDelta, JWT, and 2-1 (APC 1.1 and APC 2.1) | 3 | Patient strain | Yes | Yes |
| 40 | Chronic osteomyelitis | *Bacteroides fragilis* | UZM3 | 4 | Patient strain | No | No |
| 43 | Lung transplant infection | *Enterococcus faecium* | EFgrKN and EFgrNG | 2 | Patient strain | Yes | No |
| 46 | Disseminated bronchiectasis | *Staphylococcus aureus*<br>*Stenotrophomonas maltophilia* | ISP<br>BUCT700 | 6<br>2 | Patient strain | Yes | No |
| 55 | Prosthetic knee infection | *Staphylococcus epidermidis* | ISP* | 4 | ATCC6538 | No | No |
| 66 | Cystic fibrosis lung infection | *Mycobacterium abscessus* | 8UZL | 5 | Patient strain | No | No |
| 82 | Cystic fibrosis lung infection | *Pseudomonas aeruginosa* | 4P and DP1 | 3 | 573 | Yes | Yes |

\**Staphylococcus aureus* bacteriophage ISP was pre-adapted (using 4 serial passages), on five strains, from five different patients to better target *Staphylococcus epidermidis* strains.

**Extended Data Table 4.** Reported adverse events in the 100 consecutive bacteriophage therapy cases.

| Patient number | Description | Bacteriophage product(s) | Infection type(s) | Application route(s) | Duration | Severity | Relationship to BT | Action | Patient outcome |
| --- | --- | --- | --- | --- | --- | --- | --- | --- | --- |
| 3 | Pulmonary septic shock | BFC 1 | Lower respiratory tract infection | Nebulization | NA | Severe | Unlikely | BT stopped | Died |
| 11 | Coughing after product inhalation* | BFC 2 | Lower respiratory tract infection | Nebulization and oral | 6 days | Moderate | Possible | None | Recovered |
| 20 | Abdominal discomfort* | BFC 1 | Abdominal and bloodstream infection | Intralesional and intravenous | 2 days | Mild | Possible | BT interrupted | Recovered |
| 31 | Rash lips* | ISP | Ear-nose-throat infection | Nasal spray | 1 day | Moderate | Possible | BT stopped | Recovered |
| 39 | Fever* | ISP | Bone infection | Intralesional | 1 day | Mild | Possible | None | Recovered |
| 42 | Local redness and pain* | PyoPhage (Eliava) | Bone infection | Intralesional | 1 day | Moderate | Possible | None | Recovered |
| 44 | Septic shock | M1 | Lower respiratory tract and urinary tract infection | Nebulization and bladder rinsing | NA | Severe | Unlikely | BT stopped | Died |
| 58 | Heart failure | ISP | Diabetic foot infection | Topical | NA | Severe | Unlikely | BT stopped | Recovered |
| 69 | Cardiogenic shock | M1 | Abdominal infection | Intralesional | NA | Severe | Unlikely | BT stopped | Died |
| 79 | Postoperative ileus | PNM and PT07 | Chronic spondylodiscitis | Intravenous | NA | Severe | Unlikely | BT stopped | Died |
| 88 | Slight increase in temperature* | E4 and Efs7 | Bone infection | Intra-articular and intravenous | Transient | Mild | Possible | None | Recovered |
| 93 | Tumor progression | 14-1 | Empyema and spinocellular carcinoma | Intralesional | NA | Severe | Unlikely | BT stopped | Died |
| 96 | Septic shock | 14-1, PNM and PT07 | Burn wound infection and blood stream infection | Topical | NA | Severe | Unlikely | BT stopped | Died |
| 99 | Diarrhea and abdominal pain* | ISP | Chronic sinusitis | Nasal spray | 25 days | Moderate | Possible | Dedicated medication | Recovered |
| 100 | Nausea | ISP | Surgical wound infection with fistula | Intralesional | 1 day | Mild | Unlikely | Dedicated medication | Recovered |

BT, bacteriophage therapy; NA, not applicable. \*Considered to be adverse reactions (a causal relationship between BT and an event was suspected and was reported).

**Extended Data Fig. 1.**
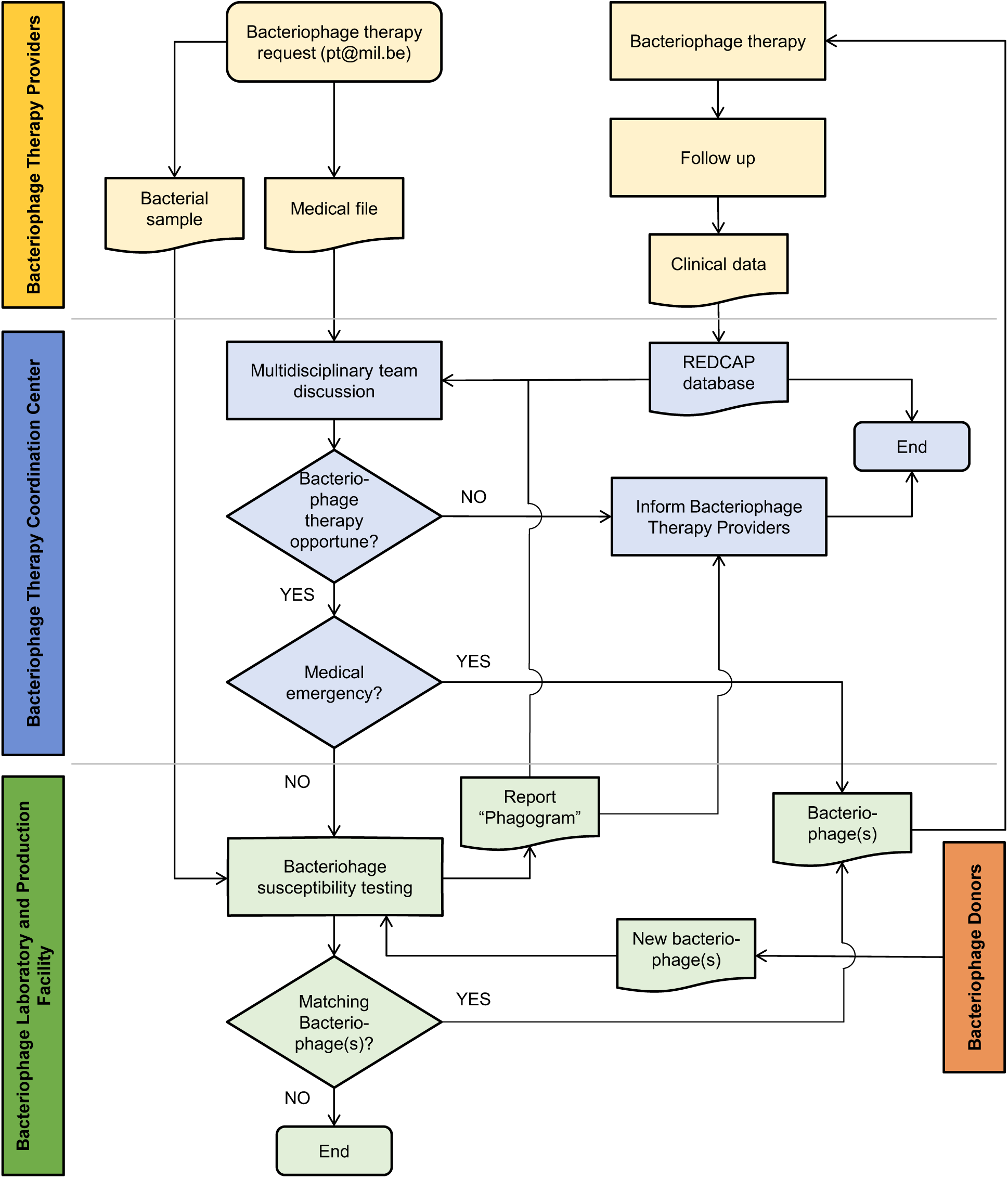
The Phage Therapy Coordination Centre’s patient selection process for bacteriophage therapy.

**Extended Data Fig. 2.**
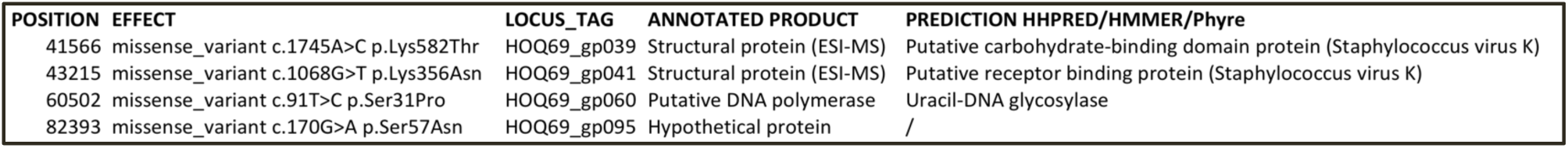
Missense mutations in the pre-adapted variant of bacteriophage ISP, as compared to the original clone (before adaptation). HHpred (https://www.sciencedirect.com/science/article/pii/S0022283617305879), HMMR (https://nar.oxfordjournals.org/content/46/W1/W200), and Phyre (https://www.nature.com/articles/nprot.2009.2) were used for functional prediction. ESI-MS, electrospray ionization mass spectrometry.

**Extended Data Fig. 3.**
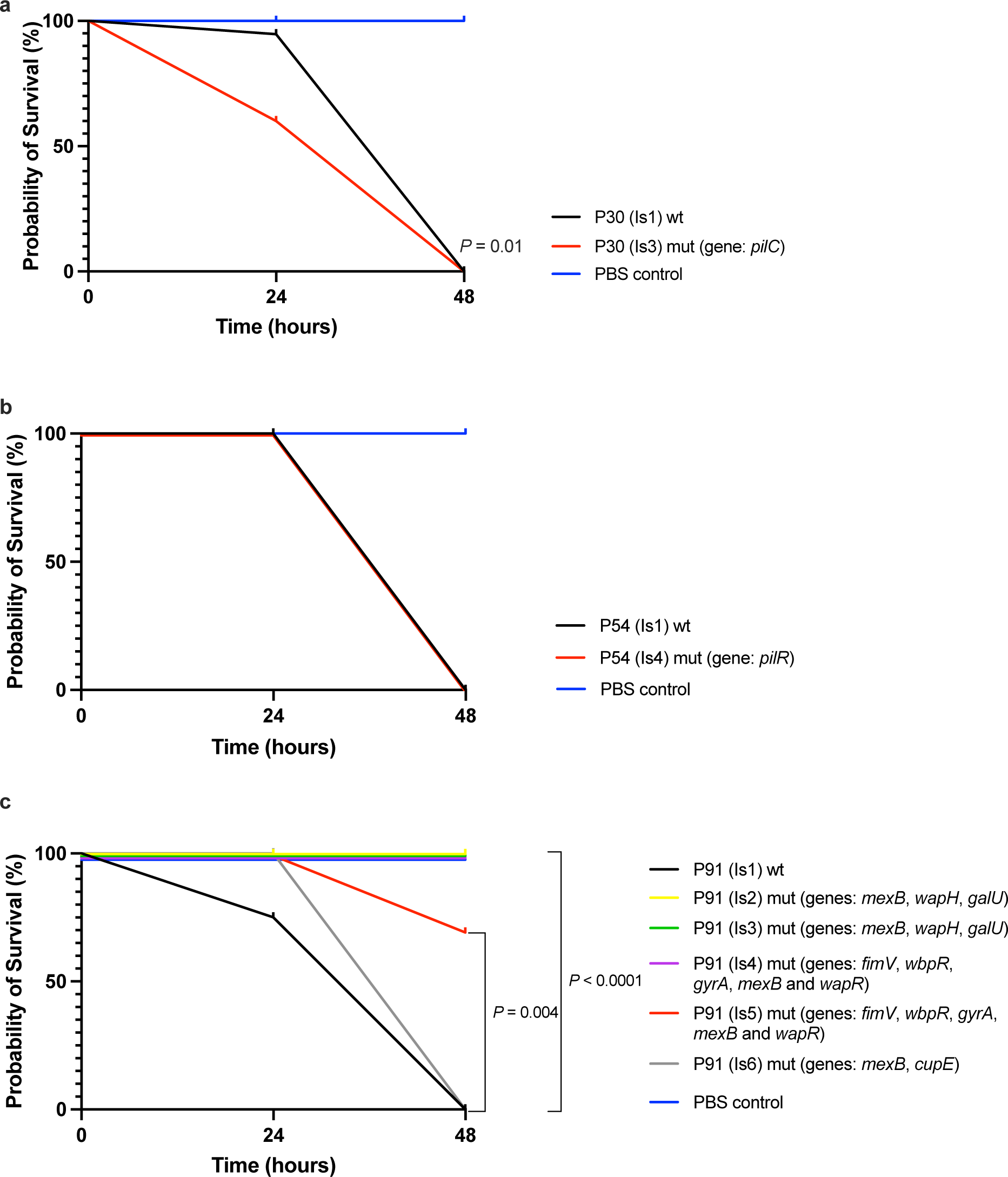
Kaplan-Meier plots showing percentage survival of *Galleria mellonella* larvae over 48 hours. Ten larvae in each group were either inoculated with phosphate buffered saline (PBS, control), with the initial bacteriophage- susceptible isolates (wild type, wt), or with the in vivo selected bacteriophage-insensitive mutants of the *Pseudomonas aeruginosa* strains isolated from patient (P) 91, 54, and 30. **a**, P30 (Is1) and P30 (Is3). **b**, P54 (Is1) and P54 (Is4). **c**, P91 (Is1-6). *P* values were calculated using the log-rank test with Bonferroni correction for multiple comparisons. Is, isolate; mut, mutation.

**Extended Data Fig. 4.**
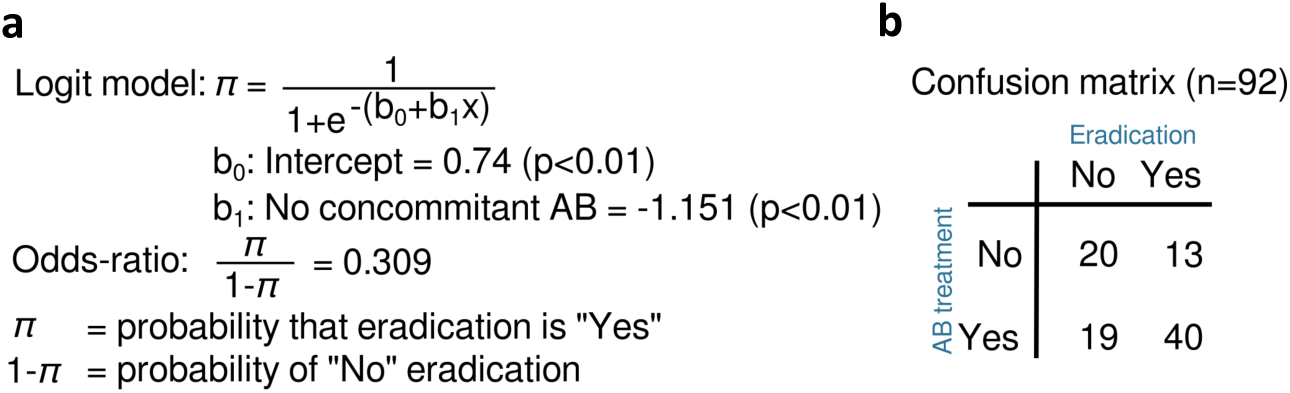
Sketch (a) and confusion matrix (b) of the logistic regression model used to analyze the reduced data set (Supplementary Table 2). AB, antibiotic.

**Extended Data Fig. 5.**
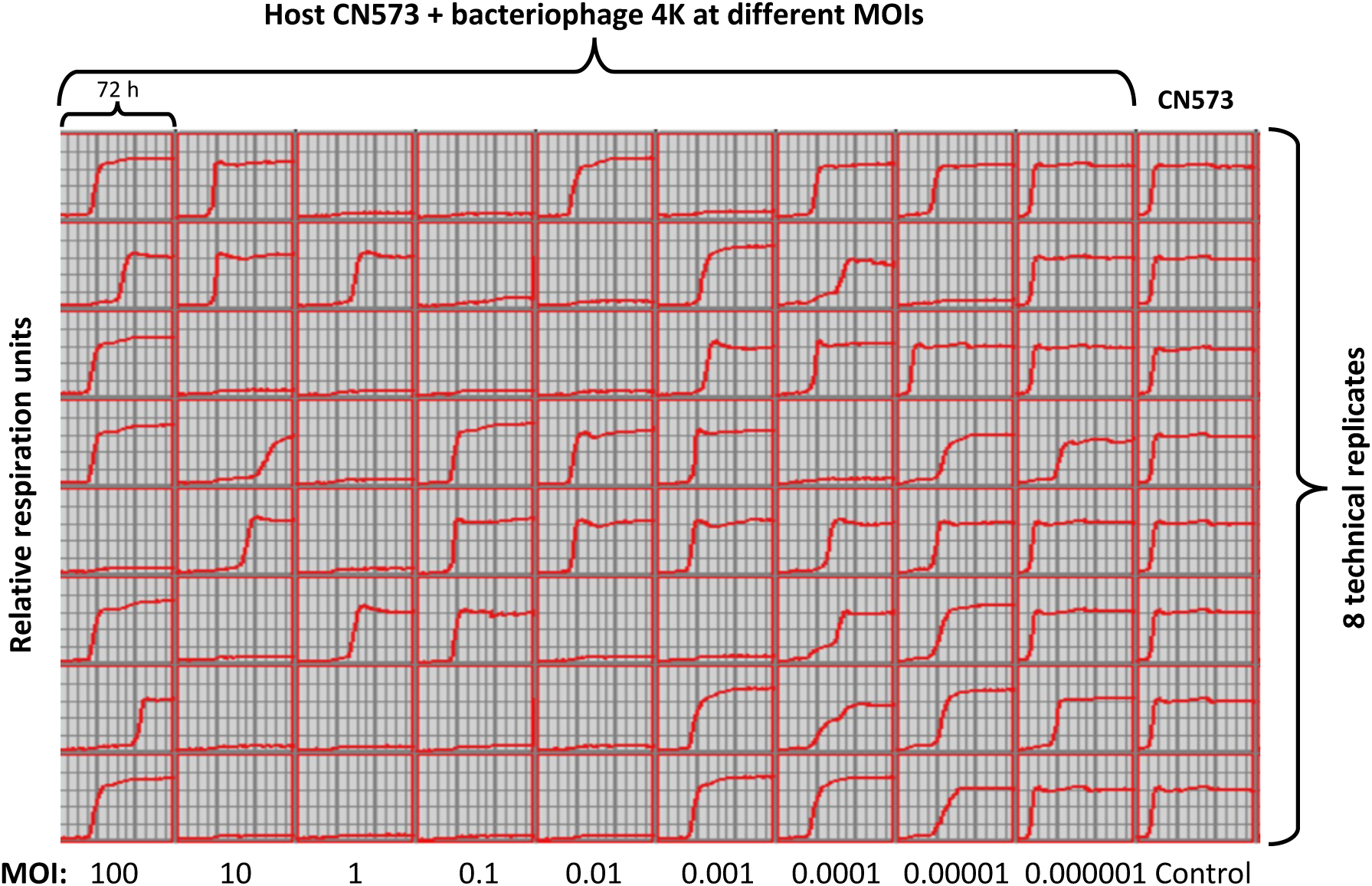
Results of the in vitro evaluation of the influence of serial multiplicities of infection (MOIs) on the virulence and on the resistance suppression of *Pseudomonas aeruginosa* bacteriophage 4K on the bacterial host strain CN573, as determined in liquid culture, using an OmniLog^®^ system. Bacterial proliferation is presented through relative units of cellular respiration over time (72 hours).

## References

1. Antimicrobial Resistance Collaborators. Global burden of bacterial antimicrobial resistance in 2019: a systematic analysis. Lancet 399, 629–655 (2022).

2. McCall, B. New fund stimulates the ailing antibiotic pipeline. Lancet Infect. Dis. 20, 1017 (2020).

3. Dublanchet, A. & Fruciano, E. Brève histoire de la phagothérapie [A short history of phage therapy]. Med. Mal. Infect. 38, 415–420 (in French) (2008).

4. Thiel, K. Old dogma, new tricks—21st Century phage therapy. Nat. Biotechnol. 22, 31–36 (2004).

5. Uyttebroek, S. et al. Safety and efficacy of phage therapy in difficult-to-treat infections: a systematic review. Lancet Infect. Dis. 22, e208–e220 (2022).

6. Pirnay, J.-P. & Kutter, E. Bacteriophages: it’s a medicine, Jim, but not as we know it. Lancet Infect. Dis. 21, 309–311 (2021).

7. Schooley, R. T. et al. Development and Use of Personalized Bacteriophage-Based Therapeutic Cocktails to Treat a Patient with a Disseminated Resistant *Acinetobacter baumannii* Infection. Antimicrob. Agents Chemother. 61, e00954–17 (2017).

8. Eskenazi, A. et al. Combination of pre-adapted and antibiotics for treatment of fracture-related infection due to pandrug-resistant *Klebsiella pneumoniae*. Nat. Commun. 13, 302 (2022).

9. Dedrick, R. M. et al. Engineered bacteriophages for treatment of a patient with a disseminated drug-resistant *Mycobacterium abscessus*. Nat. Med. 25, 730–733 (2019).

10. Pirnay, J.-P. et al. The phage therapy paradigm: prêt-à-porter or sur-mesure? Pharm. Res. 28, 934–937 (2011).

11. Verbeken, G. & Pirnay, J.-P. European regulatory aspects of phage therapy: magistral phage preparations. Curr. Opin. Virol. 52, 24–29 (2022).

12. Aslam, S. et al. Lessons Learned from the First 10 Consecutive Cases of Intravenous Bacteriophage Therapy to Treat Multidrug-Resistant Bacterial Infections at a Single Center in the United States. Open Forum Infect. Dis. 7, ofaa389 (2020).

13. Petrovic Fabijan, A., et al. Safety of bacteriophage therapy in severe *Staphylococcus aureus* infection. Nat. Microbiol. 5, 465–472 (2020).

14. Ferry, T. et al. Phage Therapy as Adjuvant to Conservative Surgery and Antibiotics to Salvage Patients with Relapsing *S. aureus* Prosthetic Knee Infection. Front. Med. (Lausanne) 7, 570572 (2020).

15. Dedrick, R. M. et al. Phage Therapy of *Mycobacterium* Infections: Compassionate-use of Phages in Twenty Patients with Drug-Resistant Mycobacterial Disease. Clin. Infect. Dis. 76, 103–112 (2022).

16. Young, M. J. et al. Phage Therapy for Diabetic Foot Infection: A Case Series. Clin. Ther. 11, S0149–2918(23)00202-3 (2023).

17. Onallah, H., Hazan, R., Nir-Paz, R., Israeli Phage Therapy Center (IPTC) Study Team. Compassionate Use of Bacteriophages for Failed Persistent Infections During the First 5 Years of the Israeli Phage Therapy Center. Open Forum Infect. Dis. 10, ofad221 (2023).

18. Onallah, H. et al. Refractory *Pseudomonas aeruginosa* infections treated with phage PASA16: A compassionate use case series. Med. S2666-6340(23)00225-8 (2023).

19. Pirnay, J. P., Verbeken, G., Ceyssens, P. J., Huys, I., De Vos, D., Ameloot, C., Fauconnier, A. The Magistral Phage. Viruses 10, 64 (2018).

20. Onsea, J. et al. Bacteriophage Therapy for Difficult-to-Treat Infections: The Implementation of a Multidisciplinary Phage Task Force (The PHAGEFORCE Study Protocol). Viruses 13, 1543 (2021).

21. Ministry of Health of the USSR and Ministry of Medical and Microbiological Industry of the USSR. Instructions for the administration of liquid staphylococcal bacteriophage preparations for injection (in Russian) (1987).

22. Ministry of Health of the USSR and Ministry of Medical and Microbiology industry of the USSR. Instructions for the application of liquid streptococcal bacteriophage preparations (in Russian) (1987).

23. Ministry of Medical and Microbiology Industry of the USSR. Instructions for the application of combined liquid pyobacteriophage preparations (in Russian) (1989).

24. Vogt, D. et al. “Beyond antibiotic therapy” – Zukünftige antiinfektiöse Strategien – Update 2017 [Beyond antibiotic therapy - Future antiinfective strategies - Update 2017]. Unfallchirurg 120, 573–584 (in German) (2017).

25. Jennes, S. et al. Use of bacteriophages in the treatment of colistin-only-sensitive *Pseudomonas aeruginosa* septicaemia in a patient with acute kidney injury-a case report. Crit. Care 21, 129 (2017).

26. Lebeaux, D. et al. A Case of Phage Therapy against Pandrug-Resistant *Achromobacter xylosoxidans* in a 12-Year-Old Lung-Transplanted Cystic Fibrosis Patient. Viruses 13, 60 (2021).

27. Van Nieuwenhuyse, B. et al. Bacteriophage-antibiotic combination therapy against extensively drug-resistant *Pseudomonas aeruginosa* infection to allow liver transplantation in a toddler. Nat. Commun. 13, 5725 (2022).

28. Van Nieuwenhuyse, B. et al. A Case of In Situ Phage Therapy against *Staphylococcus aureus* in a Bone Allograft Polymicrobial Biofilm Infection: Outcomes and Phage-Antibiotic Interactions. Viruses 13, 1898 (2021).

29. Onsea, J. et al. Bacteriophage Application for Difficult-to-treat Musculoskeletal Infections: Development of a Standardized Multidisciplinary Treatment Protocol. Viruses 11, 891 (2019).

30. Ferry, T. et al. Personalized bacteriophage therapy to treat pandrug-resistant spinal *Pseudomonas aeruginosa* infection. Nat. Commun. 13, 4239 (2022).

31. Racenis, K. et al. Use of Phage Cocktail BFC 1.10 in Combination with Ceftazidime-Avibactam in the Treatment of Multidrug-Resistant *Pseudomonas aeruginosa* Femur Osteomyelitis - A Case Report. Front. Med. (Lausanne) 9, 851310 (2022).

32. Bakuradze, N. et al. Characterization of a Bacteriophage GEC_vB_Bfr_UZM3 Active against *Bacteroides fragilis*. Viruses 15, 1042 (2023).

33. Paul, K. et al. Bacteriophage Rescue Therapy of a Vancomycin-Resistant *Enterococcus faecium* Infection in a One-Year-Old Child following a Third Liver Transplantation. Viruses 13, 1785 (2021).

34. Tkhilaishvili, T. et al. Successful case of adjunctive intravenous bacteriophage therapy to treat left ventricular assist device infection. J. Infect. 83, e1–e3 (2021).

35. Racenis, K. et al. Successful Bacteriophage-Antibiotic Combination Therapy against Multidrug-Resistant *Pseudomonas aeruginosa* Left Ventricular Assist Device Driveline Infection. Viruses 15, 1210 (2023).

36. Blasco, L. et al. Case report: Analysis of phage therapy failure in a patient with a *Pseudomonas aeruginosa* prosthetic vascular graft infection. Front. Med. 10, 1199657 (2023).

37. Takeuchi, I. et al. The Presence of Two Receptor-Binding Proteins Contributes to the Wide Host Range of Staphylococcal Twort-Like Phages. Appl. Environ. Microbiol. 82, 5763–5774 (2016).

38. Sergueev, K. V. et al. Correlation of Host Range Expansion of Therapeutic Bacteriophage Sb-1 with Allele State at a Hypervariable Repeat Locus. Appl. Environ. Microbiol. 85, e01209–e01219 (2019).

39. Treepong, P. et al. Global emergence of the widespread *Pseudomonas aeruginosa* ST235 clone. Clin. Microbiol. Infect. 24, 258–266 (2018).

40. Hrabák, J. et al. Regional spread of *Pseudomonas aeruginosa* ST357 producing IMP-7 metallo-β-lactamase in Central Europe. J. Clin. Microbiol. 49, 474–475 (2011).

41. Kilmury, S. L. N. & Burrows, L. L. The *Pseudomonas aeruginosa* PilSR Two-Component System Regulates Both Twitching and Swimming Motilities. mBio 9, e01310–e01318 (2018).

42. Nunn, D., Bergman, S. & Lory, S. Products of three accessory genes, pilB, pilC, and pilD, are required for biogenesis of *Pseudomonas aeruginosa* pili. J. Bacteriol. 172, 2911–2919 (1990).

43. Wehbi, H. et al. The peptidoglycan-binding protein FimV promotes assembly of the *Pseudomonas aeruginosa* type IV pilus secretin. J. Bacteriol. 193, 540–550 (2011).

44. Kropinski, A. M., Chan, L., Jarrell, K. & Milazzo, F. H. The nature of *Pseudomonas aeruginosa* strain PAO bacteriophage receptors. Can. J. Microbiol. 23, 653–658 (1977).

45. Koderi Valappil, S., et al. Survival Comes at a Cost: A Coevolution of Phage and Its Host Leads to Phage Resistance and Antibiotic Sensitivity of *Pseudomonas aeruginosa* Multidrug Resistant Strains. Front. Microbiol. 12, 783722 (2021).

46. Yoshida, H., Bogaki, M., Nakamura, M. & Nakamura, S. Quinolone resistance-determining region in the DNA gyrase *gyrA* gene of *Escherichia coli*. Antimicrob. Agents Chemother. 34, 1271–1272 (1990).

47. Takenouchi, T., Sakagawa, E. & Sugawara, M. Detection of *gyrA* mutations among 335 *Pseudomonas aeruginosa* strains isolated in Japan and their susceptibilities to fluoroquinolones. Antimicrob. Agents Chemother. 43, 406–409 (1999).

48. Yonezawa, M. et al. Analysis of the NH2-terminal 87^th^ amino acid of *Escherichia coli* GyrA in quinolone-resistance. Microbiol. Immunol. 39, 517–520 (1995).

49. Nakajima, A., Sugimoto, Y., Yoneyama, H. & Nakae, T. High-level fluoroquinolone resistance in *Pseudomonas aeruginosa* due to interplay of the MexAB-OprM efflux pump and the DNA gyrase mutation. Microbiol. Immunol. 46, 391–395 (2002).

50. Chanishvili, N. A literature review of the practical application of bacteriophage research (Nova Biomedical Books, 2012).

51. Verbeken, G. et al. Taking bacteriophage therapy seriously: a moral argument. Biomed. Res. Int. 2014, 621316 (2014).

52. Church, D., Elsayed, S., Reid, O., Winston, B. & Lindsay, R. Burn wound infections. Clin. Microbiol. Rev. 19, 403–434 (2006).

53. Rose, T. et al. Experimental phage therapy of burn wound infection: difficult first steps. Int. J. Burns Trauma 4, 66–73 (2014).

54. Friman, V. P. et al. Pre-adapting parasitic phages to a pathogen leads to increased pathogen clearance and lowered resistance evolution with *Pseudomonas aeruginosa* cystic fibrosis bacterial isolates. J. Evol. Biol. 29, 188–198 (2016).

55. Appelmans, R. Le dosage du Bacteriophage. Compt. Rend. Soc. Biol. 85, 1098 (in French) (1921).

56. Burrowes, B. H., Molineux, I. J. & Fralick, J. A. Directed in Vitro Evolution of Therapeutic Bacteriophages: The Appelmans Protocol. Viruses 11, 241 (2019).

57. Pirnay, J.-P. Phage Therapy in the Year 2035. Front. Microbiol. 11, 1171 (2020).

58. Cano, E.J., et al. Phage Therapy for Limb-threatening Prosthetic Knee *Klebsiella pneumoniae* Infection: Case Report and In Vitro Characterization of Anti-biofilm Activity. Clin. Infect. Dis. 73, e144–e151 (2021).

59. Suh, G. A. et al. Considerations for the Use of Phage Therapy in Clinical Practice. Antimicrob. Agents Chemother. 66, e0207121 (2022).

60. Labrie, S. J., Samson, J. E. & Moineau, S. Bacteriophage resistance mechanisms. Nat. Rev. Microbiol. 8, 317– 327 (2010).

61. Luria, S. E. & Delbrück, M. Mutations of Bacteria from Virus Sensitivity to Virus Resistance. Genetics 28, 491–511 (1943).

62. Oechslin, F. et al. Synergistic Interaction Between Phage Therapy and Antibiotics Clears Pseudomonas Aeruginosa Infection in Endocarditis and Reduces Virulence. J. Infect. Dis. 215, 703–712 (2017).

63. Castledine, M. et al. Parallel evolution of *Pseudomonas aeruginosa* phage resistance and virulence loss in response to phage treatment *in vivo* and *in vitro*. Elife 11, e73679 (2022).

64. Westra, E.R. et al. Parasite exposure drives selective evolution of constitutive versus inducible defense. Curr. Biol. 25,1043–1049 (2015).

65. Gu Liu, C., et al. Phage-Antibiotic Synergy Is Driven by a Unique Combination of Antibacterial Mechanism of Action and Stoichiometry. mBio 11, e01462–20 (2020).

66. Fungo, G. B. N. et al. “Two Is Better Than One”: The Multifactorial Nature of Phage-Antibiotic Combinatorial Treatments Against ESKAPE-Induced Infections. Phage (New Rochelle) 4, 55–67 (2023).

67. Torres-Barceló, C. & Hochberg, M. E. Evolutionary Rationale for Phages as Complements of Antibiotics. Trends Microbiol. 24, 249–256 (2016).

68. Torres-Barceló, C. Phage Therapy Faces Evolutionary Challenges. Viruses 10, 323 (2018).

69. Chan, B. K., Sistrom, M., Wertz, J. E., Kortright, K. E., Narayan, D., Turner, P. E. Phage selection restores antibiotic sensitivity in MDR *Pseudomonas aeruginosa*. Sci. Rep. 6, 26717 (2016).

70. Abedon, S. T. Phage-Antibiotic Combination Treatments: Antagonistic Impacts of Antibiotics on the Pharmacodynamics of Phage Therapy? Antibiotics 8, 182 (2019).

71. Becker, Y. Replication of Viral and Cellular Genomes: Molecular Events at the Origins of Replication and Biosynthesis of Viral and Cellular Genomes (Springer, 1983).

72. Górski, A. et al. Phage as a modulator of immune responses: practical implications for phage therapy. Adv. Virus Res. 83, 41–71 (2012).

73. Ministry of Health of the USSR. Instructions for the application of a liquid staphylococcal phage preparation for injection (in Russian) (1986).

74. Łusiak-Szelachowska, M. et al. Phage neutralization by sera of patients receiving phage therapy. Viral Immunol. 27, 295–304 (2014).

75. Gembara, K. & Dąbrowska, K. Phage-specific antibodies. Curr. Opin. Biotechnol. 68, 186–192 (2021).

76. Dedrick, R. M. et al. Potent antibody-mediated neutralization limits bacteriophage treatment of a pulmonary *Mycobacterium abscessus* infection. Nat. Med. 27, 1357–1361 (2021).

77. Hopewell, S., Loudon, K., Clarke, M. J., Oxman, A. D. & Dickersin, K. Publication bias in clinical trials due to statistical significance or direction of trial results. Cochrane Database Syst. Rev. 2009, MR000006 (2009).

## References

78. REGULATION (EU) No 536/2014 OF THE EUROPEAN PARLIAMENT AND OF THE COUNCIL of 16 April 2014 on clinical trials on medicinal products for human use, and repealing Directive 2001/20/EC. 2014. http://ec.europa.eu/health//sites/health/files/files/eudralex/vol-1/reg_2014_536/reg_2014_536_en.pdf.

79. Merabishvili, M. et al. Quality-controlled small-scale production of a well-defined bacteriophage cocktail for use in human clinical trials. PLoS One 4, e4944 (2009).

80. Bolger, A. M., Lohse, M. & Usadel, B. Trimmomatic: a flexible trimmer for Illumina sequence data. Bioinformatics 30, 2114–2120 (2014).

81. Wick, R. R., Judd, L. M., Gorrie, C. L. & Holt, K. E. Unicycler: Resolving bacterial genome assemblies from short and long sequencing reads. PLoS Comput. Biol. 13, e1005595 (2017).

82. Prjibelski, A., Antipov, D., Meleshko, D., Lapidus, A. & Korobeynikov, A. Using SPAdes De Novo Assembler. Curr. Protoc. Bioinformatics. 70, e102 (2020).

83. Seemann, T. Prokka: rapid prokaryotic genome annotation. Bioinformatics 30, 2068–2069 (2014).

84. Arndt, D. et al. PHASTER: a better, faster version of the PHAST phage search tool. Nucleic Acids Res. 44, W16–W21 (2016).

85. Song, W. et al. Prophage Hunter: an integrative hunting tool for active prophages. Nucleic Acids Res. 47, W74–W80 (2019).

86. Kutter, E. Phage host range and efficiency of plating. Methods Mol. Biol. 501, 141–149 (2009).

87. Ministry of Health of the USSR. Department for industrial bacterial and viral preparations. Guidelines for the production of combined pyobacteriophage solutions. N242-82 (in Russian) (1982).

88. Ministry of Health of the USSR. Guidelines for the production of staphylococcal bacteriophage solutions for injection (in Russian) (1986).

89. Ministry of Health of the USSR. Pharmacopoeia Commission. Department for the monitoring of the introduction of new medicines and medical equipment. Staphylococcal bacteriophage solutions for injection. BФC 42-68BC-87 (in Russian) (1987).

90. Ministry of Health of the USSR. Pharmacopoeia commission. Pharmacopoeia article concerning combined pyobacteriophage solutions. ФC 42-240BC-8 (in Russian) (1989).

91. Merabishvili, M., Pirnay, J.-P. & De Vos, D. Guidelines to Compose an Ideal Bacteriophage Cocktail. Methods Mol. Biol. 1693, 99–110 (2018).

92. Duyvejonck, H. et al. Evaluation of the Stability of Bacteriophages in Different Solutions Suitable for the Production of Magistral Preparations in Belgium. Viruses 13, 865 (2021).

93. Merabishvili, M. et al. Stability of bacteriophages in burn wound care products. PLoS One 12, e0182121 (2017).

94. Astudillo, A., Leung, S. S. Y., Kutter, E., Morales, S. & Chan, H. K. Nebulization effects on structural stability of bacteriophage PEV 44. Eur. J. Pharm. Biopharm. 125, 124–130 (2018).

95. Carrigy, N. B. et al. Anti-Tuberculosis Bacteriophage D29 Delivery with a Vibrating Mesh Nebulizer, Jet Nebulizer, and Soft Mist Inhaler. Pharm. Res. 34, 2084–2096 (2017).

96. Cantalapiedra, C. P., Hernández-Plaza, A., Letunic, I., Bork, P. & Huerta-Cepas, J. eggNOG-mapper v2: Functional Annotation, Orthology Assignments, and Domain Prediction at the Metagenomic Scale. Mol. Biol. Evol. 38, 5825–5829 (2021).

97. Brown, C. L. et al. mobileOG-db: a Manually Curated Database of Protein Families Mediating the Life Cycle of Bacterial Mobile Genetic Elements. Appl. Environ. Microbiol. 88, e0099122 (2022).

98. Starikova, E. V. et al. Phigaro: high-throughput prophage sequence annotation. Bioinformatics 36, 3882–3884 (2020).

99. Krzywinski, M. et al. Circos: an information aesthetic for comparative genomics. Genome Res. 19, 1639– 1645 (2009).

100. Gao, F. & Zhang, C. T. GC-Profile: a web-based tool for visualizing and analyzing the variation of GC content in genomic sequences. Nucleic Acids Res. 34, W686–W691 (2006).

101. Page, A. J. et al. Roary: rapid large-scale prokaryote pan genome analysis. Bioinformatics 31, 3691–3693 (2015).

102. Letunic, I. & Bork, P. Interactive Tree Of Life (iTOL) v5: an online tool for phylogenetic tree display and annotation. Nucleic Acids Res. 49, W293–W296 (2021).

103. Adams, M. H. Bacteriophages (Interscience Publishers, 1959).

104. Harris, P. A. et al. Research electronic data capture (REDCap) – A metadata-driven methodology and workflow process for providing translational research informatics support. J. Biomed. Inform. 42, 377–381 (2009).

105. Magiorakos, A. P. et al. Multidrug-resistant, extensively drug-resistant and pandrug-resistant bacteria: an international expert proposal for interim standard definitions for acquired resistance. Clin. Microbiol. Infect. 18, 268–281 (2012).

106. McDonnell, A. et al. Efficient Delivery of Investigational Antibacterial Agents via Sustainable Clinical Trial Networks. Clin. Infect. Dis. 63, S57–S59 (2016).

107. R Core Team. R: A language and Environment for Statistical Computing (R Foundation for Statistical Computing, 2016).

108. Wickham, H. et al. Welcome to the tidyverse. JOSS 4, 1686 (2019).

109. Conway, J. R., Lex, A. & Gehlenborg, N. UpSetR: an R package for the visualization of intersecting sets and their properties. Bioinformatics 33, 2938–2940 (2017).

110. Kahle, D. & Wickham, H. ggmap: Spatial Visualization with ggplot2. The R Journal 5, 144–161 (2013).

111. Massicotte, P. & South, A. rnaturalearth: World Map Data from Natural Earth (rnaturalearth, 2023).

